# Distinct Neurocognitive Profiles and Clinical Phenotypes Associated with Copy Number Variation at the 22q11.2 Locus

**DOI:** 10.1101/2023.05.12.23289905

**Authors:** Kathleen P. O’Hora, Leila Kushan-Wells, Gil D. Hoftman, Maria Jalbrzikowski, Raquel C. Gur, Ruben Gur, Carrie E. Bearden

## Abstract

Rare genetic variants that confer large effects on neurodevelopment and behavioral phenotypes can reveal novel gene-brain-behavior relationships relevant to autism. Copy number variation at the 22q11.2 locus offer one compelling example, as both the 22q11.2 deletion (22qDel) and duplication (22qDup) confer increased likelihood of autism spectrum disorders (ASD) and cognitive deficits, but only 22qDel confers increased psychosis risk. Here, we used the Penn Computerized Neurocognitive Battery (Penn-CNB) to characterized neurocognitive profiles of 126 individuals: 55 22qDel carriers (M_Age_=19.2 years, 49.1% male), 30 22qDup carriers (M_Age_=17.3 years, 53.3 % male), and 41 typically developing (TD) subjects (M_Age_=17.3 years, 39.0 % male). We performed linear mixed models to assess group differences in overall neurocognitive profiles, domain scores, and individual test scores. We found all three groups exhibited distinct overall neurocognitive profiles. 22qDel and 22qDup carriers showed significant accuracy deficits across all domains relative to controls (Episodic Memory, Executive Function, Complex Cognition, Social Cognition, and Sensorimotor Speed), with 22qDel carriers exhibiting more severe accuracy deficits, particularly in Episodic Memory. However, 22qDup carriers generally showed greater slowing than 22qDel carriers. Notably, slower social cognition speed was uniquely associated with increased global psychopathology and poorer psychosocial functioning in 22qDup. Compared to TD, 22q11.2 CNV carriers failed to show age-associated improvements in multiple cognitive domains. Exploratory analyses revealed 22q11.2 CNV carriers with ASD exhibited differential neurocognitive profiles, based on 22q11.2 copy number. These results suggest that there are distinct neurocognitive profiles associated with either a loss or gain of genomic material at the 22q11.2 locus.

## Introduction

Neurocognitive profiles are of clinical importance for individuals with neurodevelopmental disorders, as cognitive performance has been shown to relate to severity of communication and social difficulties (Demetriou et al., 2018; Leung, Vogan, Powell, Anagnostou, & Taylor, 2016; Oliveras-Rentas, Kenworthy, Roberson, Martin, & Wallace, 2012) and repetitive behaviors (Mostert-Kerckhoffs, Staal, Houben, & de Jonge, 2015), can differentiate individuals with Autism Spectrum Disorders (ASD) (Doi, Kanai, & Ohta, 2022; Nader, Jelenic, & Soulières, 2015; Solomon et al., 2021), potentially predict behavioral challenges, and is often used to aid diagnoses (Nader et al., 2015). As such, classifying individuals based on neurocognitive profiles could provide a framework for uncovering genetic and neurobiological differences associated with autism phenotypes, allowing for improved diagnosis and clinical outcomes. However, because of the phenotypic diversity within the behaviorally-defined (idiopathic) autism spectrum, identifying neurocognitive profiles associated with specific autism phenotypes requires large sample sizes, which is challenging to accomplish due to the special needs of the autism population. An alternative, potentially more feasible strategy is to study individuals with rare genetic variants that confer large effects on neurodevelopment and behavioral phenotypes related to autism. Deep phenotyping of individuals with these rare variants can reveal genetic, neurobiological, and psychological mechanisms that influence developmental processes in autism. Copy Number Variants (CNVs), genetic variants in which relatively large sections of the genome are duplicated or deleted, in genomic regions influencing neurodevelopment greatly increase the likelihood of autism and are present in up to 11% of individuals with autism (Devlin & Scherer, 2012; Geschwind, 2011; Mahjani et al., 2021). Because the molecular basis of these highly-penetrant CNVs are known, they offer a powerful method for examining relationships between genes, brain, and behaviors, which can be used to understand typical neurodevelopment and reveal shared biological mechanisms with idiopathic autism (Moreau, Ching, Kumar, Jacquemont, & Bearden, 2021). Supporting this notion, several studies have identified common neurobiological and genetic mechanisms in CNVs and idiopathic autism (Connacher et al., 2022; Moreau et al., 2021).

CNVs in the 22q11.2 locus, a genomic hotspot for neurodevelopmental genes (Guna, Butcher, & Bassett, 2015), differentially modulate cognitive and clinical phenotypes (Lin et al., 2017; Olsen et al., 2018), making them a strong model to investigate specific neurocognitive profiles associated with autism and autism-related phenotypes. A 1.5 to 2.6 megabase hemizygous deletion at the 22q11.2 locus (22qDel) is the most common contiguous gene deletion syndrome, occurring in approximately 1 in 4000 live births (McDonald-McGinn et al., 2015). It is associated with congenital malformations, developmental delay and intellectual disability, as well as multiple neurodevelopmental disorders, including psychotic spectrum disorder, ASD, attention deficit hyperactivity disorder (ADHD) and anxiety disorders (Schneider et al., 2014). Although less well-characterized, duplications in the 22q11.2 region (22qDup) are ∼2.5 times more common than 22q11.2 deletions (Olsen et al., 2018). A 22qDup also confers increased likelihood of autism (Olsen et al., 2018; Verbesselt, Zink, Breckpot, & Swillen, 2022) as well as ADHD and cognitive impairment. Recent research has identified intriguing gene-dosage effects of the 22q11.2 locus on brain structure (Lin et al., 2017), clinical symptoms (Lin et al., 2020), speech-language and cognition (Lin et al., 2020; Verbesselt et al., 2023). Interestingly, the two CNVs confer similar likelihood of autism and similar deficits in social cognition but, they differentially modulate intellectual ability and psychosis-risk (Lin et al., 2020; Rees et al., 2014). Both CNVs are associated with lower Intelligence Quotient (IQ) scores but, consistent with the generally more deleterious effects of a deletion relative to a duplication (Männik et al., 2015), the 22qDel confers a larger impact on IQ than the 22qDup (Lin et al., 2020). The differential impact of the 22q11.2 CNVs on intellectual ability, along with their convergence on autism risk and impaired social cognition, make the study of specific neurocognitive profiles of 22q11.2 CNV carriers a critical step towards a mechanistic understanding of cognitive phenotypes relevant to autism.

Previous examinations of neurocognitive profiles of individuals with a 22qDel identified widespread deficits in neurocognitive performance, with notable impairments in executive function, episodic memory, and social cognition (Goldenberg et al., 2012; R. E. Gur et al., 2014; Moberg et al., 2018; Yi et al., 2016). Cognition is impaired from a young age and develops at a slower rate in 22qDel carriers relative to TD youth (R. E. Gur et al., 2014; Vorstman et al., 2015). In comparison, little is known about the neurocognitive profiles of 22qDup carriers. There are a handful of reports of decreased intellectual functioning in 22qDup carriers (Alberti et al., 2007; Chawner, Owen, et al., 2019; Olsen et al., 2018; Verbesselt et al., 2022) and studies in mouse models suggest that overexpression of 22q11.2 genes influences working memory throughout development via dysregulated hippocampal neurogenesis (Boku et al., 2018; Suzuki et al., 2009). However, to our knowledge, a recent report from our research group is the only published study examining specific domains of cognitive performance in 22qDup carriers (Lin et al., 2020); this study found deficits in working memory, verbal memory, processing speed, and social cognition in 22qDup carriers compared to typically developing (TD) subjects (Lin et al., 2020). These deficits were similar in magnitude to those observed in 22qDel carriers. However, 22qDup carriers scored higher than 22qDel, but lower than TD subjects, on measures of general cognition (nonverbal, verbal, and full-scale IQ).

Both global cognitive impairments and impairments in specific neurocognitive processes have been shown to be related to psychosis, general psychopathology, and social functioning in 22qDel carriers (Antshel, Fremont, Ramanathan, & Kates, 2017; Antshel et al., 2010; Chawner, Niarchou, et al., 2019; Gothelf, Penniman, Gu, Eliez, & Reiss, 2007; Gothelf et al., 2013; Jalal et al., 2021; Morrison et al., 2020; Vorstman et al., 2015; Weinberger et al., 2016). 22qDel carriers with psychosis exhibit greater cognitive impairment than 22qDel carriers without psychosis (Antshel et al., 2010; Morrison et al., 2020; Yi et al., 2016) and a steeper decline in general cognition (i.e. IQ) is associated with increased subsequent psychosis risk in 22qDel carriers (Gothelf et al., 2007, 2013; Vorstman et al., 2015). Interestingly, emergence of psychosis in 22qDel carriers was associated with worsening performance on attention-executive tasks (Antshel et al., 2017; Chawner, Niarchou, et al., 2019), which predicted psychosis emergence better than measures of global cognition (Chawner, Niarchou, et al., 2019). In terms of social functioning, within adolescents with 22qDel, autism-related behaviors were associated with poorer processing speed, sustained attention, and working memory (Morrison et al., 2020). Further, social cognition performance was positively correlated with verbal IQ in 22qDel (Jalal et al., 2021; Lin et al., 2020). The one study to date to investigate this in reciprocal 22q11.2 CNV carriers found a similar pattern in 22qDup, but there was a stronger correlation in 22qDup carriers (Lin et al., 2020). Taken together, this evidence suggests investigation of cognitive measures beyond IQ in 22q11.2 CNV carriers may reveal unique differences in neurocognitive profiles, which could better predict social function and clinical outcomes.

The present study seeks to expand on previous work investigating cognition in 22q11.2 CNV carriers through use of a rigorous, well-validated computerized cognitive battery to characterize differences in neurocognitive profiles between 22qDel carriers, 22qDup carriers, and TD controls. We hypothesize that 22q11.2 CNV carriers exhibit distinct neurocognitive profiles, which can be used to distinguish individuals with a 22qDel from those with a 22qDup. Specifically, we hypothesize 22qDel carriers will exhibit more profound neurocognitive deficits in executive control, complex cognition, and episodic memory compared to 22qDup carriers. In contrast, we expect the two CNV groups to show similar levels of impairment in social cognition. Next, we aimed to determine if cognitive deficits observed in 22qDel carriers are related to differential clinical outcomes between the two CNV groups. We hypothesize that poorer cognitive performance will be associated with more severe overall psychopathology and positive symptoms, as well as poorer psychosocial functioning in the 22qDel group, but not the 22qDup group. In both CNV groups, we expect poorer social cognition to be associated with worse real-world social functioning. Lastly, we conducted exploratory analyses examining neurocognitive profiles of individuals with autism within each CNV group to investigate how autism may differentially influence neurocognition in 22q11.2 CNV carriers.

## Methods

### Participants

The total sample consisted of 126 subjects: 55 subjects with a molecularly confirmed 22q11.2 deletion, 30 subjects with a molecularly confirmed 22q11.2 duplication, and 41 demographically comparable TD subjects. Participants were a subset of those recruited for an ongoing longitudinal study conducted at University of California at Los Angeles (UCLA). Patients were recruited from local clinics, national support groups, and other online avenues. Participants for the community-control sample were recruited from the same communities as CNV carriers through online postings, local schools, pediatric clinics, and other locations in the community. Subjects returned for two follow-up visits, each one year apart. While data acquisition is still ongoing at the time of analysis, all data available to date are included in the current analysis (208 total datapoints across 126 subjects). A subset of participants (60%) in this study were included in a previous analysis (Lin et al., 2020). The previous investigation included cross-sectional measures of global cognition derived from the Wechsler Abbreviated Scale of Intelligence (D. Wechsler, 1999) (WASI) and Wechsler Adult Intelligence Scale (David Wechsler, n.d.) (WAIS) as well as two Penn Computerized Neurocognitive Battery (R. C. Gur et al., 2010; Moore, Reise, Gur, Hakonarson, & Gur, 2015) (Penn-CNB) subtests (Emotion Recognition and Emotion Differentiation). The present analysis expands upon the previous report by including more comprehensive neurocognitive measures from the Penn-CNB, including separate accuracy and speed scores, and including 51 additional subjects and 82 longitudinal datapoints.

Participants with significant neurological or medical conditions (unrelated to 22q11.2 CNVs) affecting brain structure or function, previous head trauma with loss of consciousness, insufficient English fluency, and/or substance abuse/dependence within the past six months were excluded. All participants gave verbal and written informed consent to participate in the study. Participants under 18 years of age provided written assent and their parent/guardian provided written consent. The UCLA Institutional Review Board approved all study procedures and documents.

### Assessments

#### Penn Neurocognitive Battery (Penn-CNB)

We employed an online neurocognitive battery that has been extensively validated across a wide age range and assesses multiple domains of cognition, including executive function and attention, verbal and nonverbal memory, verbal and non-verbal reasoning, social cognition, and sensorimotor speed (R. C. Gur et al., 2010; Moore et al., 2015). The battery has been used in children with intellectual disability(Yi et al., 2016), youth at clinical high risk for psychosis (Goldenberg et al., 2012), and with genetic disorders (O’Hora, Zhang, et al., 2022), including 22qDel carriers (Goldenberg et al., 2012; R. C. Gur et al., 2021; Weinberger et al., 2016). It captures accuracy and speed measures and employs automated quality assurance and scoring procedures (Moore et al., 2015). A detailed description of the Penn-CNB tasks is reported in prior publications (Goldenberg et al., 2012; R. C. Gur et al., 2012, 2010; Moore et al., 2015). Penn-CNB domains and subtests are summarized in Table 1.

**Table 1.** Summary of Cognition Measures

| General Cognition | Domain | Test |
| --- | --- | --- |
| Global<br>Neurocognitive<br>Performance (GNP) | Executive Control | Mental Flexibility (ABF) |
|  |  | Attention (ATT) |
|  |  | Working Memory (WM) |
|  | Episodic Memory | Verbal Memory (VMEM) |
|  |  | Face Memory (FMEM) |
|  |  | Spatial Memory (SMEM) |
|  | Complex Cognition | Verbal Reasoning (VR) |
|  |  | Nonverbal reasoning (NVR) |
|  |  | Spatial Ability (SPA) |
|  | Social Cognition | Emotion Identification (EI) |
|  |  | Emotion Differentiation (ED) |
|  |  | Age Differentiation (AD) |
|  | Sensorimotor | Motor Praxis (MOT) |
|  |  | Sensorimotor (SM) |

Accuracy and speed (reaction time for correct trials) values for each test were z-transformed to the TD group in the current sample. All speed scores were multiplied by −1 so that a lower z-score represents worse (slower) performance. For outliers, z-scores <-4 were set to a floor value of −4. A global neurocognitive performance (GNP) measure was calculated by taking the mean of the z-scores across all Penn-CNB tests.

#### Structured Interview for Psychosis-risk Syndromes (SIPS)

The SIPS (Miller et al., 2004), a clinician-rated semi-structured interview, was used to measure positive psychosis-risk symptoms. The measure uses information from both the participant and their parent to determine clinical ratings. Participants over the age of ten completed the SIPS interview, administered by a trained clinician. The primary outcome was positive symptom score.

#### Brief Psychiatric Rating Scale (BPRS)

The total score on the BPRS (Overall & Gorham, 1962), a well-validated and highly reliable clinician-rated scale that assesses 24 items across several domains of psychopathology (Roncone et al., 1999), was used to measure global psychopathology.

#### Global Assessment of Functioning Scale (GAF)

The GAF (Hall, 1995) is an overall measure of current psychiatric well-being that ranges from positive mental health to severe psychopathology. It is a generic scale that is not diagnosis-specific and has previously been used in 22q11.2 CNV carriers (Lin et al., 2020).

#### Child Behavior Checklist (CBCL)

The Social Problems scale on the CBCL (Achenbach, 1999; Achenbach & Rescorla, 2001) was used as a measure of real-world social functioning. The CBCL is a well-validated parent-report scale used as a dimensional measure of psychopathology in children and adolescents. Individual T-scores based on the population norm of the Social Problems subscale were used in the present analyses.

#### Autism Assessment

Participants were assessed for autism spectrum disorder diagnosis by a trained clinician using the Autism Diagnostic Interview-Revised (ADI-R) (Lord, Rutter, & Le Couteur, 1994) and the Autism Diagnostic Observation Schedule 2^nd^ Edition (ADOS-2) (Gotham, Risi, Pickles, & Lord, 2006; Lord, Rutter, Di Lavore, & Risi, 2012). The ADOS and ADI-R are gold standard diagnostic assessments for autism. The ADOS relies on a qualified examiner’s direct observation of interaction abilities and behaviors while the ADI-R interviews a parent or caretaker about the individual’s developmental history and current behavior.

### Data analysis

All analyses were conducted in R version 4.0.2 (R Core Team, 2021) using the packages nlme (Pinheiro, Bates, & R Core Team, 2022) and MASS (Venables & Ripley, 2002). Figures were made using ggplot2 (Wickham, 2016). All analyses included all available data, which includes up to three timepoints from each subject.

First, we assessed group differences in global cognition (GNP) across the three subject groups using linear mixed models (LMM) with GNP as the outcome variable and subject group as the predictor variable. Age (in months) and sex were included as covariates in the model and subject ID was included as the random effects term.

Next, we performed two types of analyses to test the hypothesis that 22qDel and 22qDup carriers show distinct neurocognitive profiles. In the first analysis, we tested for group differences in specific neurocognitive profiles between 22qDel, 22qDup, and TD controls, following similar procedures to those used in Service et al. 2021 (Service et al., 2020). Briefly, z-scores for each test were modeled as a function of subject group (with TD as the reference group), sex, age, cognitive test (14 individual measures in Table 1), and all interactions using LMMs, with individual Penn-CNB tests as repeated measures. If a significant interaction of subject group and Penn-CNB test was observed, we performed separate LMMs to test for pairwise differences in Penn-CNB domain scores (5 domains in Table 1) between groups (22qDel vs. TD, 22qDup vs. TD, 22qDel vs. 22qDup), with subject ID as the random effects term. For domain scores with significant main effects of group, we assessed pairwise group differences in the individual tests composing that domain. Multiple comparison correction using False Discovery Rate (FDR) was applied to account for the number of cognitive measures assessed in this analysis. As in Service et al 2021 (Service et al., 2020), separate analyses were done for accuracy and speed due to differing factorial structures (Moore et al., 2015). As a secondary analysis to determine whether anti-psychotic medication use contributing to differences in neurocognitive profiles, we separately included this variable in the LMM. Exploratory analyses were conducted in subgroup of participants with autism diagnostic data available (n=40 22qDel, n=28 22qDup; Sample characteristics in Table S1) to examine neurocognitive profiles of autistic individuals within each CNV group. We aimed to investigate whether these individuals will exhibit unique neurocognitive phenotypes, depending on 22q11.2 gene-dosage. For these exploratory analyses, subjects were classified into four groups based on CNV type and ASD diagnosis (22qDelASD-, 22qDelASD+, 22qDupASD-, 22qASD+). Differences in overall neurocognitive profiles between the four groups were tested using the same LMM procedure outlined above. However, due to decreased statistical power and the exploratory nature of these analyses, analyses of pairwise group differences focused on Penn-CNB domain scores, rather than individual tests.

The second set of analyses examining the existence of distinct neurocognitive profiles between 22q11.2 CNV carriers investigated if neurocognitive data could be used to discriminate between 22qDel and 22qDup carriers. Fisher’s Linear Discrimination Analysis (LDA) was used to distinguish between CNV carriers based on performance on the Penn-CNB tests. We used separate models for accuracy and speed and compared model performance, which was assessed using leave-one-out cross-validation. We conducted 100 simulations and computed the mean and standard deviation of accuracies for each model. To account for the differences in sample size, the 22qDup was over-sampled. A “control” model was constructed by randomly shuffling identities within the training set. Mean accuracy of each model was compared to the control and each other to determine if one model was superior in discriminating between 22qDel and 22qDup carriers.

Lastly, to test whether differences in specific cognitive domains are related to the differential clinical phenotypes between CNV groups, we assessed the relationship between cognitive performance and psychosis symptoms (measured by the SIPS), social problems (measured by the CBCL), and global psychopathology (measured by the BPRS). We additionally assessed relationships between cognitive performance and global psychosocial function (measured by the GAF). We conducted LMMs, with 22qDel as the reference group, to test for interactions between CNV group and clinical variables for Penn-CNB domain scores. The regression coefficient of the clinical variable term was used to assess relationships between cognitive and clinical variables within the reference group. The reference group was re-leveled and the same model was run to assess relationships within the 22qDup group. FDR correction was applied within each clinical domain to correct for the number of comparisons.

## Results

There were no significant differences in age, sex, or depressive disorder diagnosis between groups, but there were significantly fewer non-white subjects in the 22qDup group compared to TD (Table 2). The 22qDup group also had significantly lower parental education compared to TD.

**Table 2.** Demographic and clinical characteristics at baseline

|  | TD | 22qDel | 22qDup |
| --- | --- | --- | --- |
| *n* | 41 | 55 | 30 |
| Age, Years (SD) | 17.3 (8.2) | 19.2 (10.8) | 17.3 (13.0) |
| Age Range, Years | 6-45 | 5-51 | 6-49 |
| Males, *n* (%) | 16 (39.0%) | 27 (49.1%) | 16 (53.3%) |
| Non-white, *n* (%)<sup>a</sup> | 10 (24.4%) | 10 (18.2%) | 1 (3.3%) |
| Highest Parental Education, Years (SD)<sup>a</sup> | 16.65 (3.6) | 15.75 (3.0) | 14.6 (2.8) |
| Psychotic Spectrum Symptoms<sup>b</sup>, *n* (%)<sup>d</sup> | 1 (2.4%) | 12 (21.8%) | 3 (10.0%) |
| Autism, *n* (%)<sup>c,e</sup> | 0 (0.0%) | 18 (32.7%) | 11 (36.7%) |
| ADHD, *n* (%)<sup>c,e</sup> | 2 (4.9%) | 25 (45.5%) | 13 (43.3%) |
| Anxiety Disorder, *n* (%)<sup>c,e</sup> | 0 (0.0%) | 24 (42.9%) | 16 (53.3%) |
| Depressive Disorder, *n* (%) | 2 (6.9%) | 8 (17.0%) | 3 (10.0%) |
| Medication, *n* (%) |  |  |  |
| Anti-Psychotics | 0 (0.0%) | 6 (10.9%) | 2 (6.7%) |
| Mood Stabilizers<sup>c</sup> | 0 (0.0%) | 3 (5.5%) | 7 (20.3%) |
| Stimulants | 1 (2.4%) | 3 (5.5%) | 1 (3.3%) |
| Other Medication | 0 (0.0%) | 4 (7.3%) | 1 (3.3%) |
| No Medication<sup>a,d</sup> | 40 (97.6%) | 39 (70.9%) | 19 (63.3%) |
| Full Scale IQ<sup>a,d,f</sup> | 115.7 (14.5) | 82.2 (13.4) | 90.6 (19.1) |

<sup>a</sup>TD>22qDup<sup>b</sup> Any positive symptom rated >2 on the SIPS<sup>c</sup>22qDup>TD<sup>d</sup>TD>22qDel<sup>e</sup>22qDel>TD<sup>f</sup>22qDup>22qDel

### 22q11.2 CNV Carriers Exhibit Distinct Neurocognitive Profiles

Both 22qDel and 22qDup carriers exhibited decreased overall cognition (GNP) compared to TD controls (*q*<0.001; Figure 1A). LMMs revealed significant interactions between subject group and cognitive test, for both accuracy and speed (*p*<0.0001), indicating distinct neurocognitive profiles across the three groups. These results remained significant when anti-psychotic use was included in the model. A summary of these results is presented in Table S2.

**Figure 1.**
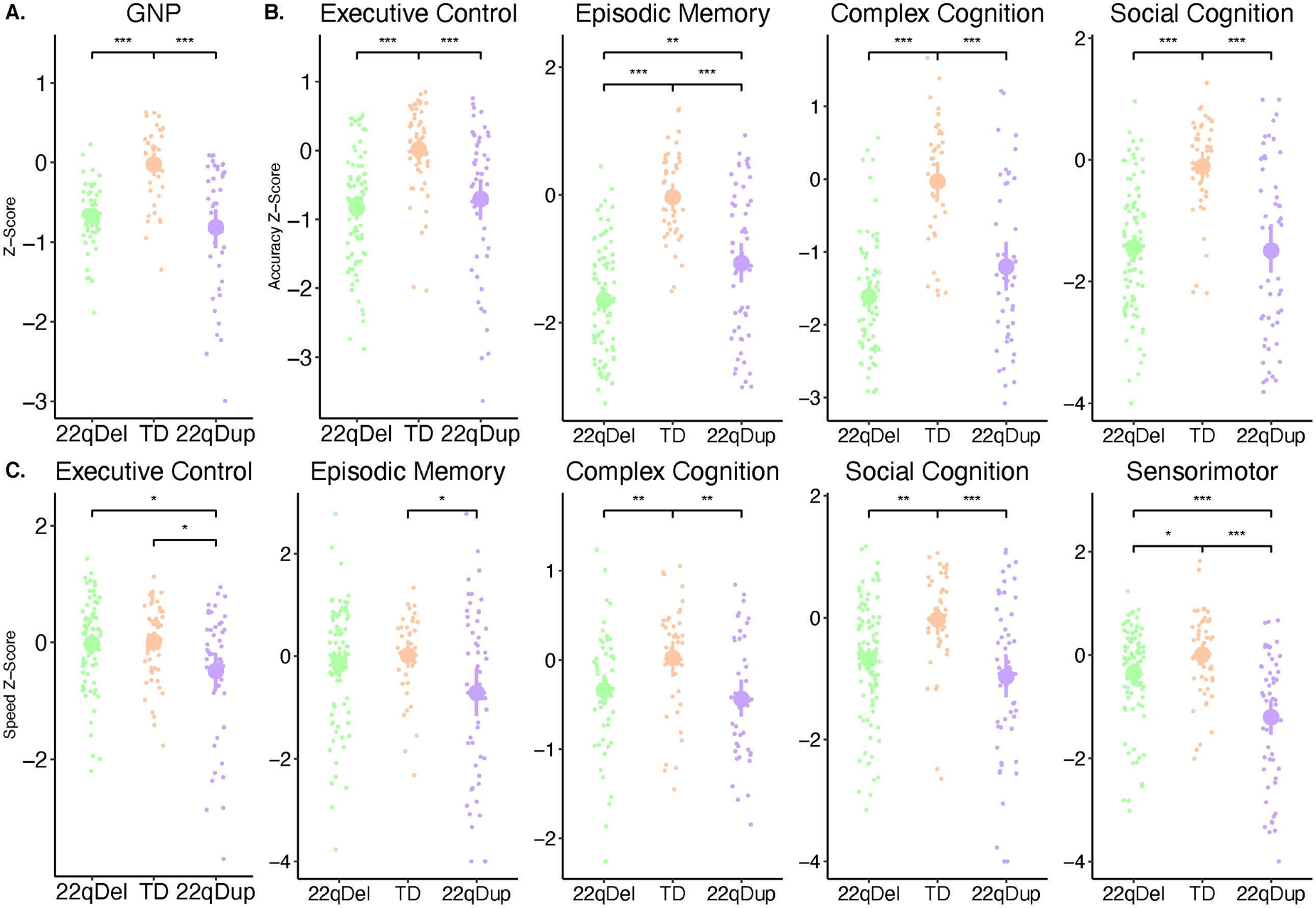
Group differences between 22qDel carriers (green), 22qDup carriers (purple), and TD controls (orange) in measures of (a) general cognition, (b) accuracy, and (c) speed for Penn-CNB neurocognitive domains (Executive Control, Episodic Memory, Complex Cognition, Social Cognition, and Sensorimotor). Large dots represent mean score and 95% bootstrapped confidence interval for each group. \**q*<0.05, \*\**q*<0.01, \*\*\**q*<0.001

Differences in Penn-CNB domain scores across the three groups are displayed in Figure 1B and 1C. There was a main effect of group on all neurocognitive domain scores (*p*<0.02). Summary of LMMs comparing Penn-CNB domain scores are presented in Table S4. Compared to TD, both CNV groups exhibited deficits in accuracy across all domains. Both CNV groups showed similar magnitudes of deficits in Social Cognition, but 22qDel carriers were less accurate than 22qDup carriers on Episodic Memory. Regarding speed, 22qDup carriers were significantly slower than TD controls in all domains, and slower than 22qDel carriers in Sensorimotor Speed and Executive Control domains. 22qDel carriers were significantly slower than controls in Sensorimotor Speed, Social Cognition, and Complex Cognition.

While we did not have a specific hypothesis regarding age interactions, we found group-by-age interactions for accuracy (*p*=0.03), but not speed (*p*=0.31), in which overall accuracy increased with age at different rates for 22qDel (*b=*0.21, *p*=0.22), 22qDup (b=0.61, *p*<0.001), and TD subjects (*b=*0.14, *p*=0.45). Conversely, there was an overall group-by-age-by-test interaction for speed, but not accuracy (Tables S2 and S3). Compared to the TD group, 22q11.2 CNV carriers showed different relationships between age and speed for emotion differentiation, mental flexibility, and age differentiation, in which the TD group’s performance improved with age, but the 22q11.2 CNV groups failed to show this age-associated improvement (Figure 2 Table S3). Similarly, the TD group performed faster with increasing age on the Verbal Reasoning test but there was no age-associated change in performance in the 22qDel group (Figure 2). Finally, there was a significant group-by-age interaction for Sensorimotor Speed between 22qDup and 22qDel carriers in which 22qDup carriers improved with age, while 22qDel carriers showed no relationship with age.

**Figure 2.**
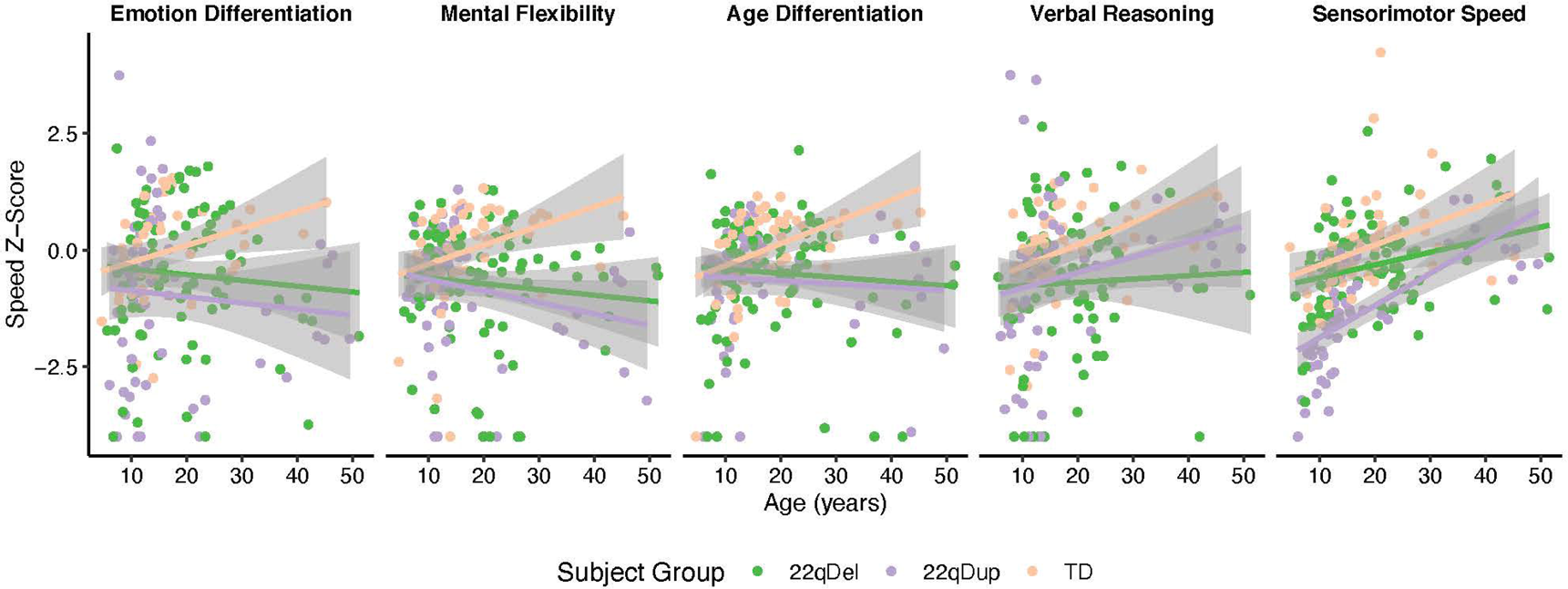
Changes in Penn-CNB Speed Scores across the age range for all subtests with a significant subject group-by-age interaction. Both CNV groups failed to show the same degree of age-related performance increases for Emotion Differentiation, Mental Flexibility, and Age Differentiation while only 22qDel carriers failed to show age-related increases for Verbal Reasoning. For Sensorimotor Speed, 22qDel carriers showed significantly less age-related increases than 22qDup carriers.

Differences in speed and accuracy for each individual Penn-CNB test are shown in Figure 3. Results of LMMs of group differences in individual test performance are summarized in Table S5. Both 22q11.2 CNV groups were less accurate than TD subjects on all Penn-CNB tests. The deficit in 22qDel carriers was more severe than that observed in 22qDup carriers for Nonverbal Reasoning and Face Memory, but the CNV groups exhibited similar deficits in accuracy relative to TD for all other subtests. 22qDup carriers were significantly slower than both 22qDel and TD groups for Attention, Sensorimotor Speed and Motor Speed. 22qDel and 22qDup carriers showed similar deficits in speed relative to TD controls on measures of Age Differentiation, Emotion Identification, and Spatial Ability. 22qDup additionally showed deficits in speed relative to TD controls in Word Memory, Emotion Differentiation, and Mental Flexibility. In contrast, 22qDel carriers were significantly faster than TD in Nonverbal Reasoning.

**Figure 3.**
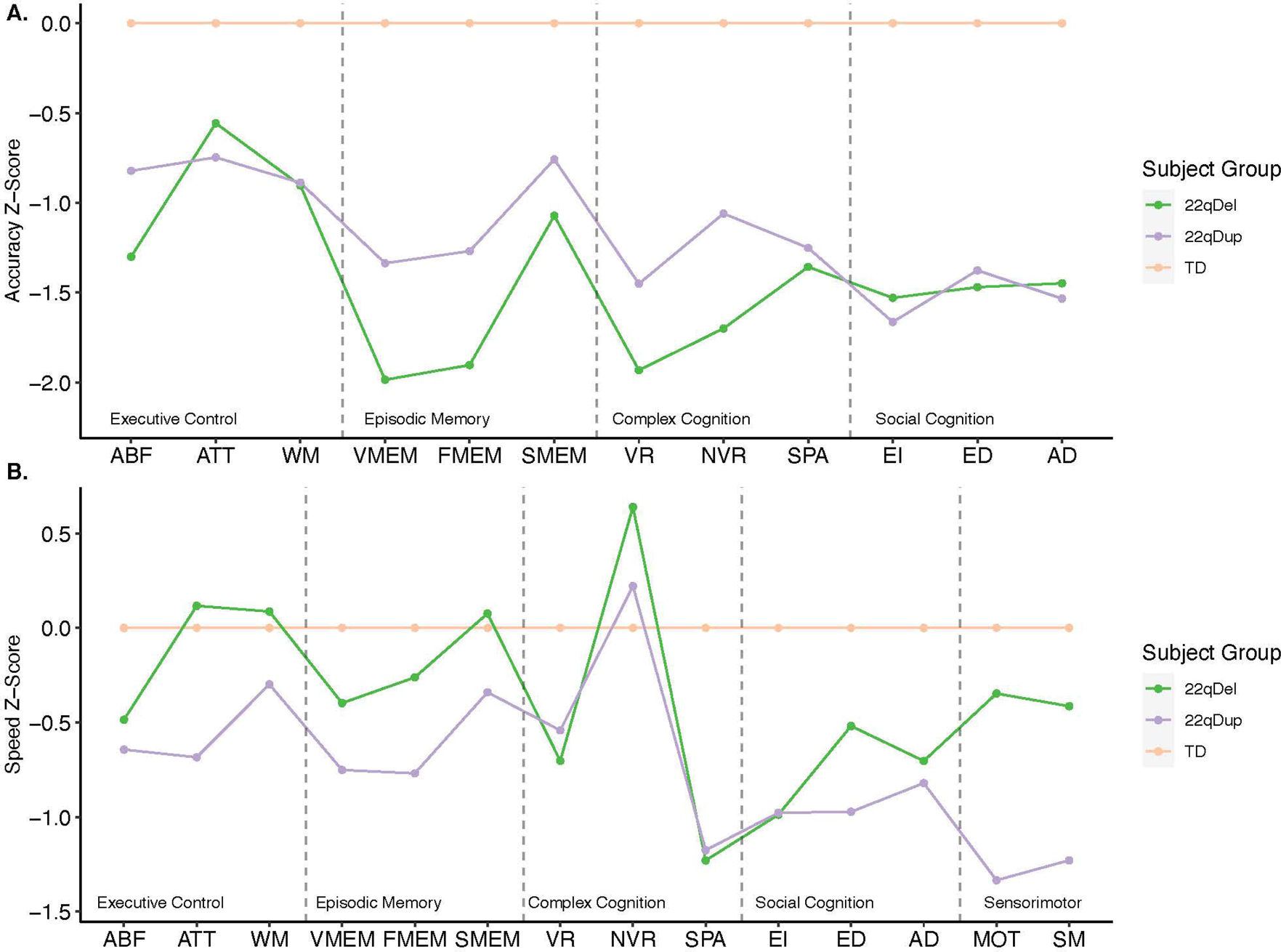
Profile of neurocognitive performance across all Penn-CNB tests in 22qDel carriers (green line), 22qDup carriers (purple line), and TD controls (orange line). Accuracy (a) and speed (b) scores are presented for mental flexibility (ABF), attention (ATT), working memory (WM), verbal memory (VMEM), face memory (FMEM), spatial memory (SMEM), verbal reasoning (VR), nonverbal reasoning (NVR), spatial processing (SPA), emotion identification (EI), emotion differentiation (ED), and age differentiation (AD). Additional speed scores are presented for motor praxis (MOT) and sensorimotor speed (SM). Both 22q CNV groups were less accurate than the TD group across all tests, with the 22qDel carriers exhibiting worse overall accuracy than 22qDup carriers. Contrastingly, 22qDup carriers exhibited worse overall speed performance than 22qDel carriers.

### Neurocognitive Profiles Successfully Discriminate between 22qDel and 22qDup Carriers

In the LDA, models successfully discriminated between 22qDel and 22qDup carriers for both accuracy (model accuracy: 76.6%, *p*<0.001) and speed (model accuracy: 83.8%, *p*<0.001). The speed model was more accurate at discriminating between the two groups (*p*=0.013) than the accuracy model (Figure S1). Within each model, the Penn-CNB tests with the largest effects in the analysis of group differences (see above) between 22qDel and 22qDups were weighted the most heavily (Accuracy: Non-Verbal Reasoning, Spatial Memory, and Face Memory; Speed: Motor Praxis, Attention, and Non-verbal Reasoning).

### Cognitive Slowing is Associated with Worse Clinical Outcomes in 22qDup Carriers, but not 22qDel Carriers

Table S6 shows a summary of the models testing the relationships between clinical measures and cognition within 22qDel and 22qDup carriers and interactions. Regression coefficients of these models are displayed in Figure 4.

**Figure 4.**
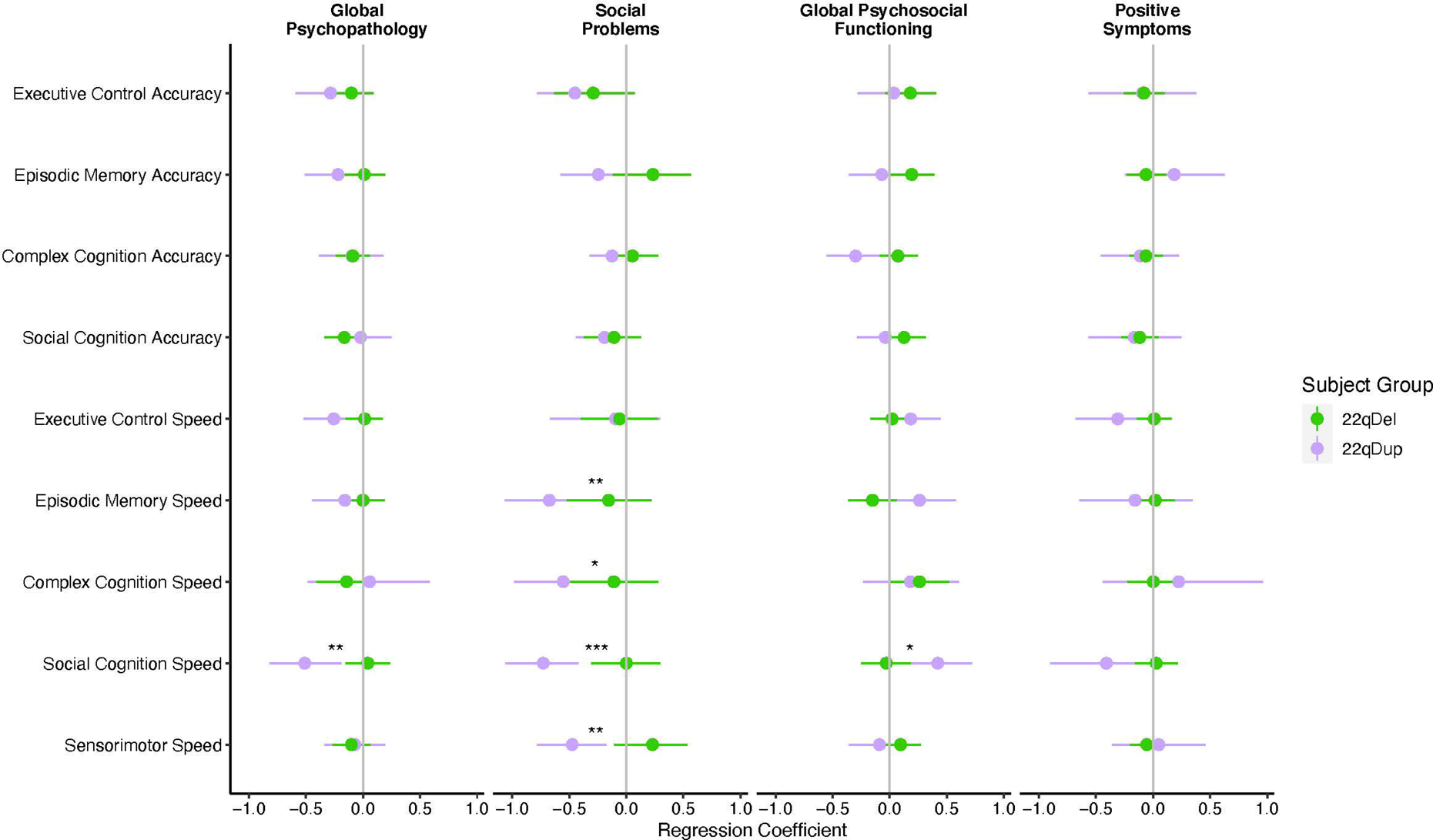
Beta coefficients and 95% confidence intervals for linear mixed models testing the association between Penn-CNB domains and clinical outcomes (Psychopathology, Social Function, Global Function, and Positive symptoms) in 22qDel carriers (green) and 22qDup carriers (purple). Worse Social Cognition speed performance was associated with increased psychopathology and worse global and social functioning in 22qDup carriers. Worse Social Functioning was also associated with worse speed performance on episodic memory, complex cognition, and sensorimotor speed domains. \**q*<0.05, \*\**q*<0.01, \*\*\**q*<0.001

There were no significant relationships between accuracy scores and clinical outcomes. There was a significant group-by-psychopathology interaction in which faster social cognition speed was associated with less severe psychopathology in 22qDup carriers, but not 22qDel carriers. Similarly, faster social cognition speed was associated with higher global functioning in 22qDup carriers, but not in 22qDel carriers. Slower speed scores on Complex Cognition, Episodic Memory, Sensorimotor, and Social Cognition were also associated with increased social problems in 22qDup carriers, but not 22qDel carriers. There was a significant group-by-social problems interaction for Sensorimotor Speed and Social Cognition Speed. In contrast, there was no relationship with speed scores and positive psychosis-risk symptoms.

### Individuals With Autism Exhibit Differential Neurocognitive Profiles, Depending on 22q11.2 Copy Number

18 (45%) 22qDel and 11 (40%) 22qDup carriers met diagnostic criteria for ASD (Tables 2, S1). LMMs revealed significantly different profiles across the four subject groups (22qDelASD-, 22qDelASD+, 22qDupASD-, and 22qDupASD+) for speed (*p*<0.001; Figure 5a), but not accuracy (*p*=0.104; Figure 5b, Table S7). Within each CNV group, there were no significant differences in Penn-CNB Domain Speed Scores between autistic individuals and their non-autistic counterparts (Figure 5c, Table S8). However, within ASD individuals, those with a 22qDup (22qDupASD+) were significantly slower on Executive Control and Sensorimotor Speed than those with a 22qDel (22qDelASD+).

**Figure 5.**
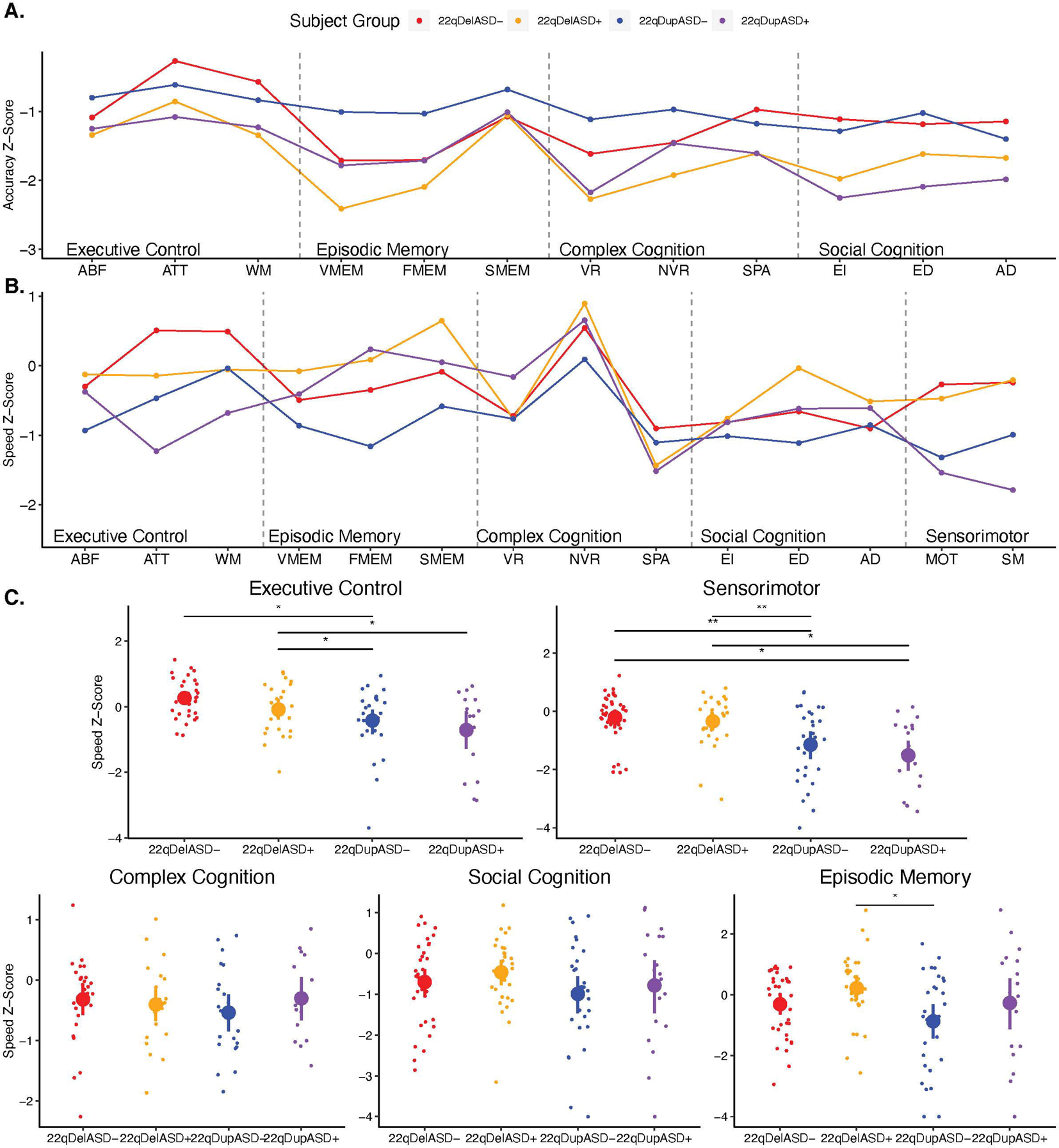
Neurocognitive Profiles of 22qDelASD-(red), 22qDelASD+ (orange), 22qDupASD-(blue), 22qASD+ (purple). Accuracy (a) and speed (b) scores are presented for for mental flexibility (ABF), attention (ATT), working memory (WM), verbal memory (VMEM), face memory (FMEM), spatial memory (SMEM), verbal reasoning (VR), nonverbal reasoning (NVR), spatial processing (SPA), emotion identification (EI), emotion differentiation (ED), and age differentiation (AD). Additional speed scores are presented for motor praxis (MOT) and sensorimotor speed (SM). LMMs revealed that distinct neurocognitive profiles exist between groups for speed (b) but not accuracy (a). (c) Group differences in speed scores for Penn-CNB neurocognitive domains (Executive Control, Episodic Memory, Complex Cognition, Social Cognition, and Sensorimotor). Large dots represent mean score and 95% bootstrapped confidence interval for each group. \**q*<0.05, \*\**q*<0.01, \*\*\**q*<0.001

## Discussion

Here, we report several novel findings regarding neurocognitive profiles in 22q11.2 CNV carriers. First, 22q11.2 CNV carriers exhibit distinct overall neurocognitive profiles, which can be used to distinguish individuals with a 22qDel from individuals with a 22qDup. Next, we observed a general accuracy versus speed trade-off in which 22qDup carriers exhibited greater accuracy, but performed slower, than 22qDel carriers. Because of this trade-off, speed scores best distinguished between the CNV groups. We also found several relationships between social cognition and clinical outcomes in 22qDup, but not 22qDel, carriers. We also found several notable group-by-age interactions in which 22q11.2 CNV carriers exhibited slower age-related improvements in tests of social cognition, verbal reasoning, and mental flexibility, relative to TD controls. Lastly,22q11.2 CNV carriers with autism exhibit differential neurocognitive profiles, based on CNV type.

As we hypothesized, there are distinct neurocognitive profiles among 22q11.2 CNV carriers, suggesting that, while both 22qDels and 22qDups exhibit global cognitive deficits, gene-dosage at the 22q11.2 locus results in differing cognitive phenotypes. A previous investigation of global cognition in 22q11.2 CNV carriers indicate that the loss of genetic material result in more cognitive impairment than gain of genetic material (Lin et al., 2020), but our findings suggest a more complex association. To our knowledge, our report of greater deficits in processing speed in 22qDup carriers is the first to suggest increased impairment in a cognitive domain in 22qDup carriers compared to 22qDel carriers. This underscores the importance of a deep phenotyping approach to characterizing more granular phenotypes, as opposed to global phenotypes, associated with rare variants.

Our analyses support and extend prior accounts of global cognitive deficits in 22q11.2 CNV carriers (Lin et al., 2020; Männik et al., 2015). But, contrary to our hypothesis, the 22qDel group scored similarly to 22qDup carriers in global cognition, measured by GNP, despite IQ differences. This is the first study to characterize differences in neurocognitive profiles between reciprocal 22q11.2 CNV carriers. 22qDup carriers exhibited deficits in accuracy across all Penn-CNB Domains relative to TD controls, but these deficits were less severe than those exhibited in 22qDel carriers, particularly for Episodic Memory. However, 22qDup carriers showed more slowing than 22qDel carriers. In particular, 22qDup carriers performed slower than 22qDel carriers in Sensorimotor and Executive Control domains. Interestingly, compared to 22qDup carriers, 22qDel carriers were both less accurate, and faster on the Non-Verbal Reasoning test. This result, along with the overall pattern of deficits in the 22q11.2 CNV carriers, suggests that there is an accuracy-speed trade-off in which 22qDup carriers sacrificed speed for increased accuracy performance. In contrast, 22qDel carriers were less accurate, but faster, which is consistent with prior accounts of Penn-CNB performance in 22qDel carriers (Goldenberg et al., 2012; R. E. Gur et al., 2014). In an independent sample of 22qDel patients, Gur et al. (2014) (R. E. Gur et al., 2014) similarly reported that Non-Verbal Reasoning was associated with faster, but less accurate responses in 22qDel carriers (R. E. Gur et al., 2014). The authors suggested this pattern reflects “giving up” during more complex, difficult tasks. However, our results suggest this phenomenon is specific to 22qDel carriers and is not exclusive to complex reasoning tasks. For example, 22qDel carriers exhibited large deficits in accuracy for tasks related to Episodic Memory, compared to both TD and 22qDup, but they scored similarly to the TD group and marginally better than 22qDup carriers in Episodic Memory Speed. However, in 22qDup carriers, accuracy deficits were mirrored by similar deficits in speed, such that they exhibited deficits in both accuracy and speed across all domains. This suggest that 22qDup carriers may have been less likely to “give-up” or lose interest during cognitive tasks. This accuracy-speed trade-off is frequently reported in the ADHD literature as attributable to lapses in attention (Mulder et al., 2010). It is possible that this result can be attributed to ADHD symptoms, which are prevalent in both 22qDel and 22qDup carriers (Olsen et al., 2018). However, in the present cohort and previously studied cohort, there is no difference in prevalence or severity of ADHD symptoms between 22qDel and 22qDup carriers (Lin et al., 2020; O’Hora, Lin, Kushan-Wells, & Bearden, 2022; Olsen et al., 2018). One potential future direction to better address this question is to examine trial-by-trial variability in response speed, which is a reliable marker of attention lapses (Chang, Lenartowicz, Hellemann, Uddin, & Bearden, 2022; Leth-Steensen, Elbaz, & Douglas, 2000). Future research is required to investigate the degree to which differences in speed between 22q11.2 CNV carriers reflect differing cognitive abilities versus other factors.

We found that a model including Penn-CNB speed score discriminated between 22qDel and 22qDup CNV carriers more accurately than a model only including accuracy scores. Similarly, speed scores were predictive of global psychopathology, social problems, and psychosocial functioning in 22qDup, but not in 22qDel, carriers. Both of these findings could be explained by the aforementioned accuracy-speed trade-off, which was only observed in 22qDel carriers and is reflected in the increased variability in speed performance between the two CNV groups. As such, speed may be a better marker of CNV type, autism symptomatology, and other clinical outcomes. While speed scores yielded better classification accuracy, both models successfully discriminated between the two CNV groups above chance levels, further confirming the existence of distinct neurocognitive profiles for both accuracy and speed.

Our prior findings suggest that 22q11.2 CNVs generally converge on deficits in social cognition (Lin et al., 2020), meaning both 22qDel and 22qDup lead to downstream impairments in social cognition and increased likelihood of autism. But, 22qDel and 22qDup carriers have been found to exhibit distinct autism-related behavioral profiles (Lin et al., 2020), implying the two CNVs may influence social cognition through different mechanistic pathways. Our results support this notion in a few different ways. First, unlike other cognitive domains, both CNV groups exhibited remarkably similar magnitudes of deficit in the overall Social Cognition domain score compared to TD, supporting convergence on social domains. However, the two groups diverge on Emotion Differentiation Speed, in which only 22qDup carriers performed significantly worse than TD controls. This suggests that a 22qDup may confer a slightly different social cognition phenotype than the 22qDel. Next, we found differential relationships between Social Cognition performance and clinical variables between the two CNV groups. In 22qDup carriers, Social Cognition Speed was predicative of global psychopathology, psychosocial functioning, and social problems, but interestingly not positive symptoms. This suggests that in 22qDup carriers, some aspects of psychopathology and behavior could both be mediated by a common underlying mechanism impacting social cognition. However, in 22qDel carriers there was no relationship between Social Cognition and clinical outcomes, suggesting that unlike 22qDup carriers, mechanisms underlying psychopathology, psychosocial functioning, and social behavior in 22qDel carriers may be distinct from social cognition. It is possible that the lack of a relationship between social cognition and clinical outcomes in 22qDel carriers could be attributed to the accuracy-speed trade-off, such that the 22qDel group’s speed scores are not representative of social cognition abilities. Additionally, since only three 22qDup carriers endorsed psychotic-spectrum symptoms at baseline (Table 2), it is possible that the BPRS and GAF scores in this group were influenced by other dimensions of psychopathology (like ASD-related symptoms), which could have a stronger relationship to Social Cognition Speed.

While large-scale longitudinal data are required to fully assess the development of distinct neurocognitive functions in 22q11.2 CNV carriers, we report several group-by-age relationships. Consistent with the convergence of 22q11.2 CNVs in the Social Cognition domain, both 22qDel and 22qDup showed slower age-related increases in Age Differentiation and Emotion Differentiation compared to TD. However, only 22qDel carriers failed to show age-related increases in Verbal Reasoning and Sensorimotor Speed, providing another potential distinction in neurocognition between 22qDel and 22qDup carriers. These results support previous reports that cognitive development is slower in 22qDel carriers compared to TD controls (R. E. Gur et al., 2014; Vorstman et al., 2015). However, these results should be interpreted carefully, due to the wide age range in each group, which was not evenly distributed across the lifespan.

Lastly, exploratory analyses revealed that autistic individuals with a 22q11.2 CNV exhibited a neurocognitive phenotype more similar to individuals with the same 22q11.2 copy number than autistic individuals with differing 22q11.2 copy number. Interestingly, this pattern was observed for speed scores, but not accuracy scores. While group differences for accuracy scores did not reach statistical significance, and therefore we did not perform further analyses, plots of neurocognitive profiles of each group (Figure 5a and 5b) suggest that, contrary to speed scores, accuracy scores may be more similar in autistic individuals, regardless of copy number at 22q11.2. However, it is likely that this exploratory analysis was underpowered to detect these neurocognitive profile differences in accuracy scores. This finding of differential neurocognitive profiles between autistic 22qDel and 22qDup carriers suggests that, while both CNVs confer similarly increased likelihood of ASD, they represent different autism phenotypes, consistent our previous finding of differential ASD symptom profiles between 22q11.2 CNV groups (Lin et al., 2020). However, the fact that there were no significant differences between autistic and non-autistic within each CNV group individuals suggests neurocognitive profiles may be more indicative of underlying genotype than autism behaviors.

While we report several important new findings, there are several limitations of this study that should be considered. First, while our sample size is large for a rare disorder, it still is limited and contains unequal sample sizes between groups. As the 22qDup group was smaller than both the 22qDel and TD groups, this could have led to fewer associations reaching statistical significance and surviving FDR correction in the 22qDup group. However, since CNVs confer large effects on brain and cognitive phenotypes (Sønderby et al., 2022), large study samples are not as critical as they are in other patient populations. While a 22qDup may be protective of developing a psychotic spectrum disorder, a handful of individuals in the 22qDup sample endorsed psychosis-risk symptoms (Table 2), which likely reflects non-specific dimensions of psychopathology. Another limitation of the study sample is baseline differences in parental education and ethnicity between the groups as well as wide age ranges, which also could have impacted results.

This study serves as the first investigation of neurocognitive profiles associated with reciprocal 22q11.2 CNVs. We identified distinct neurocognitive profiles between 22qDel and 22qDup carriers, which require future study to determine relevance to the differential clinical outcomes between the two groups over time.

## Data Availability

The data that support the findings of this study are available from the corresponding author upon reasonable request.

## Acknowledgements

We are grateful to all participants, families, and staff who contributed to data collection. This work is supported by National Institute of Mental Health Grant Nos. R01 MH085953 (to CEB), and U01MH101779 (to CEB); and the Simons Foundation (SFARI Explorer Award to CEB).

## Notes

### Competing Interest Statement

The authors have declared no competing interest.

### Author Declarations

The University of California Los Angeles Institutional Review Board gave ethical approval of all study procedures and documents.

## References

Achenbach, T. M. (1999). The Child Behavior Checklist and related instruments. The Use of Psychological Testing for Treatment Planning and Outcomes Assessment, 2nd Ed., pp. 429–466. Mahwah, NJ, US: Lawrence Erlbaum Associates Publishers.

Achenbach, T. M., & Rescorla, L. A. (2001). Manual for the ASEBA school-age forms & profiles: child behavior checklist for ages 6-18, teacher’s report form, youth self-report: an integrated system of multi-informant assessment. University of Vermont, research center for children youth & families.

Alberti, A., Romano, C., Falco, M., Calì, F., Schinocca, P., Galesi, O., … Fichera, M. (2007). 1.5 Mb de novo 22q11.21 microduplication in a patient with cognitive deficits and dysmorphic facial features. Clinical Genetics, 71(2), 177–182. doi:10.1111/j.1399-0004.2007.00750.x

Antshel, K. M., Fremont, W., Ramanathan, S., & Kates, W. R. (2017). Predicting Cognition and Psychosis in Young Adults With 22q11.2 Deletion Syndrome. Schizophrenia Bulletin, 43(4), 833–842. doi:10.1093/schbul/sbw135

Antshel, K. M., Shprintzen, R., Fremont, W., Higgins, A. M., Faraone, S. V., & Kates, W. R. (2010). Cognitive and psychiatric predictors to psychosis in velocardiofacial syndrome: a 3-year follow-up study. Journal of the American Academy of Child and Adolescent Psychiatry, 49(4), 333–344. Retrieved from https://www.ncbi.nlm.nih.gov/pubmed/20410726

Boku, S., Izumi, T., Abe, S., Takahashi, T., Nishi, A., Nomaru, H., … Hiroi, N. (2018). Copy number elevation of 22q11.2 genes arrests the developmental maturation of working memory capacity and adult hippocampal neurogenesis. Molecular Psychiatry, 23(4), 985–992. doi:10.1038/mp.2017.158

Chang, S. E., Lenartowicz, A., Hellemann, G. S., Uddin, L. Q., & Bearden, C. E. (2022). Variability in cognitive task performance in early adolescence is associated with stronger between-network anticorrelation and future attention problems. Biological Psychiatry Global Open Science. doi:10.1016/j.bpsgos.2022.11.003

Chawner, S. J. R. A., Niarchou, M., Doherty, J. L., Moss, H., Owen, M. J., & van den Bree, M. B. M. (2019). The emergence of psychotic experiences in the early adolescence of 22q11.2 Deletion Syndrome. Journal of Psychiatric Research, 109, 10–17. doi:10.1016/j.jpsychires.2018.11.002

Chawner, S. J. R. A., Owen, M. J., Holmans, P., Raymond, F. L., Skuse, D., Hall, J., & van den Bree, M. B. M. (2019). Genotype-phenotype associations in children with copy number variants associated with high neuropsychiatric risk in the UK (IMAGINE-ID): a case-control cohort study. The Lancet. Psychiatry, 6(6), 493–505. doi:10.1016/S2215-0366(19)30123-3

Connacher, R., Williams, M., Prem, S., Yeung, P. L., Matteson, P., Mehta, M., … DiCicco-Bloom, E. (2022). Autism NPCs from both idiopathic and CNV 16p11.2 deletion patients exhibit dysregulation of proliferation and mitogenic responses. Stem Cell Reports, 17(6), 1380–1394. doi:10.1016/j.stemcr.2022.04.019

Demetriou, E. A., Lampit, A., Quintana, D. S., Naismith, S. L., Song, Y. J. C., Pye, J. E., … Guastella, A. J. (2018). Autism spectrum disorders: a meta-analysis of executive function. Molecular Psychiatry, 23(5), 1198–1204. doi:10.1038/mp.2017.75

Devlin, B., & Scherer, S. W. (2012). Genetic architecture in autism spectrum disorder. Current Opinion in Genetics & Development, 22(3), 229–237. doi:10.1016/j.gde.2012.03.002

Doi, H., Kanai, C., & Ohta, H. (2022). Transdiagnostic and sex differences in cognitive profiles of autism spectrum disorder and attention-deficit/hyperactivity disorder. Autism Research: Official Journal of the International Society for Autism Research, 15(6), 1130– 1141. doi:10.1002/aur.2712

Geschwind, D. H. (2011). Genetics of autism spectrum disorders. Trends in Cognitive Sciences, 15(9), 409–416. doi:10.1016/j.tics.2011.07.003

Goldenberg, P. C., Calkins, M. E., Richard, J., McDonald-McGinn, D., Zackai, E., Mitra, N., … Gur, R. E. (2012). Computerized neurocognitive profile in young people with 22q11.2 deletion syndrome compared to youths with schizophrenia and at-risk for psychosis. American Journal of Medical Genetics. Part B, Neuropsychiatric Genetics: The Official Publication of the International Society of Psychiatric Genetics, 159B(1), 87–93. doi:10.1002/ajmg.b.32005

Gotham, K., Risi, S., Pickles, A., & Lord, C. (2006). The Autism Diagnostic Observation Schedule (ADOS). Journal of Autism and Developmental Disorders.

Gothelf, D., Penniman, L., Gu, E., Eliez, S., & Reiss, A. L. (2007). Developmental trajectories of brain structure in adolescents with 22q11.2 deletion syndrome: a longitudinal study. Schizophrenia Research, 96(1–3), 72–81. doi:10.1016/j.schres.2007.07.021

Gothelf, D., Schneider, M., Green, T., Debbané, M., Frisch, A., Glaser, B., … Eliez, S. (2013). Risk factors and the evolution of psychosis in 22q11.2 deletion syndrome: a longitudinal 2-site study. Journal of the American Academy of Child and Adolescent Psychiatry, 52(11), 1192–1203.e3. doi:10.1016/j.jaac.2013.08.008

Guna, A., Butcher, N. J., & Bassett, A. S. (2015). Comparative mapping of the 22q11.2 deletion region and the potential of simple model organisms. Journal of Neurodevelopmental Disorders, 7(1). doi:10.1186/s11689-015-9113-x

Gur, R. C., Moore, T. M., Weinberger, R., Mekori-domachevsky, E., Gross, R., Emanuel, B. S., … Gothelf, D. (2021). Relationship between intelligence quotient measures and computerized neurocognitive performance in 22q11. 2 deletion syndrome. (April), 1–10. doi:10.1002/brb3.2221

Gur, R. C., Richard, J., Calkins, M. E., Chiavacci, R., Hansen, J. A., Bilker, W. B., … Gur, R. E. (2012). Age group and sex differences in performance on a computerized neurocognitive battery in children age 8-21. Neuropsychology, 26(2), 251–265. doi:10.1037/a0026712

Gur, R. C., Richard, J., Hughett, P., Calkins, M. E., Macy, L., Bilker, W. B., … Gur, R. E. (2010). A cognitive neuroscience-based computerized battery for efficient measurement of individual differences: standardization and initial construct validation. Journal of Neuroscience Methods, 187(2), 254–262. doi:10.1016/j.jneumeth.2009.11.017

Gur, R. E., Yi, J. J., McDonald-McGinn, D. M., Tang, S. X., Calkins, M. E., Whinna, D., … Gur, R. C. (2014). Neurocognitive development in 22q11.2 deletion syndrome: comparison with youth having developmental delay and medical comorbidities. Molecular Psychiatry, 19(11), 1205–1211. doi:10.1038/mp.2013.189

Hall, R. C. (1995). Global assessment of functioning. A modified scale. Psychosomatics, 36(3), 267–275. doi:10.1016/S0033-3182(95)71666-8

Jalal, R., Nair, A., Lin, A., Eckfeld, A., Kushan, L., Zinberg, J., … Bearden, C. E. (2021). Social cognition in 22q11.2 deletion syndrome and idiopathic developmental neuropsychiatric disorders. Journal of Neurodevelopmental Disorders, 13(1), 1–15. doi:10.1186/s11689-021-09363-4

Leth-Steensen, C., Elbaz, Z. K., & Douglas, V. I. (2000). Mean response times, variability, and skew in the responding of ADHD children: a response time distributional approach. Acta Psychologica, 104(2), 167–190. doi:10.1016/s0001-6918(00)00019-6

Leung, R. C., Vogan, V. M., Powell, T. L., Anagnostou, E., & Taylor, M. J. (2016). The role of executive functions in social impairment in Autism Spectrum Disorder. Child Neuropsychology: A Journal on Normal and Abnormal Development in Childhood and Adolescence, 22(3), 336–344. doi:10.1080/09297049.2015.1005066

Lin, A., Ching, C. R. K., Vajdi, A., Sun, D., Jonas, R. K., Jalbrzikowski, M., … Bearden, C. E. (2017). Mapping 22q11.2 gene dosage effects on brain morphometry. Journal of Neuroscience, 37(26), 6183–6199. doi:10.1523/JNEUROSCI.3759-16.2017

Lin, A., Vajdi, A., Kushan-Wells, L., Helleman, G., Hansen, L. P., Jonas, R. K., … Bearden, C. E. (2020). Reciprocal Copy Number Variations at 22q11.2 Produce Distinct and Convergent Neurobehavioral Impairments Relevant for Schizophrenia and Autism Spectrum Disorder. Biological Psychiatry, 88(3), 260–272. doi:10.1016/j.biopsych.2019.12.028

Lord, C., Rutter, M., Di Lavore, P. C., & Risi, S. (2012). Autism Diagnostic Observation Schedule 2nd ed. Western Psychological Services.

Lord, C., Rutter, M., & Le Couteur, A. (1994). Autism Diagnostic Interview-Revised: a revised version of a diagnostic interview for caregivers of individuals with possible pervasive developmental disorders. Journal of Autism and Developmental Disorders, 24(5), 659– 685. doi:10.1007/BF02172145

Mahjani, B., De Rubeis, S., Gustavsson Mahjani, C., Mulhern, M., Xu, X., Klei, L., … Buxbaum, J. D. (2021). Prevalence and phenotypic impact of rare potentially damaging variants in autism spectrum disorder. Molecular Autism, 12(1), 65. doi:10.1186/s13229-021-00465-3

Männik, K., Mägi, R., Macé, A., Cole, B., Guyatt, A. L., Shihab, H. A., … Reymond, A. (2015). Copy number variations and cognitive phenotypes in unselected populations. JAMA - Journal of the American Medical Association, 313(20), 2044–2054. doi:10.1001/jama.2015.4845

McDonald-McGinn, D. M., Sullivan, K. E., Marino, B., Philip, N., Swillen, A., Vorstman, J. A. S., … Bassett, A. S. (2015). 22Q11.2 Deletion Syndrome. Nature Reviews Disease Primers, 1(November). doi:10.1038/nrdp.2015.71

Miller, T. J., McGlashan, T. H., Rosen, J. L., Cadenhead, K., Cannon, T., Ventura, J., … Woods, S. W. (2004). Prodromal assessment with the structured interview for prodromal syndromes and the scale of prodromal symptoms: Predictive validity, interrater reliability, and training to reliability (Schizophrenia Bulletin (2003) 29, 4 (703-715). Schizophrenia Bulletin, 30(2), 218. doi:10.1093/oxfordjournals.schbul.a007072

Moberg, P. J., Richman, M. J., Roalf, D. R., Morse, C. L., Graefe, A. C., Brennan, L., … Gur, R. E. (2018). Neurocognitive Functioning in Patients with 22q11.2 Deletion Syndrome: A Meta-Analytic Review. Behavior Genetics, 48(4), 259–270. doi:10.1007/s10519-018-9903-5

Moore, T. M., Reise, S. P., Gur, R. E., Hakonarson, H., & Gur, R. C. (2015). Psychometric properties of the Penn Computerized Neurocognitive Battery. Neuropsychology, 29(2), 235–246. doi:10.1037/neu0000093

Moreau, C. A., Ching, C. R., Kumar, K., Jacquemont, S., & Bearden, C. E. (2021). Structural and functional brain alterations revealed by neuroimaging in CNV carriers. Current Opinion in Genetics & Development, 68, 88–98. doi:10.1016/j.gde.2021.03.002

Morrison, S., Chawner, S. J. R. A., van Amelsvoort, T. A. M. J., Swillen, A., Vingerhoets, C., Vergaelen, E., … van den Bree, M. B. M. (2020). Cognitive deficits in childhood, adolescence and adulthood in 22q11.2 deletion syndrome and association with psychopathology. Translational Psychiatry, 10(1), 53. doi:10.1038/s41398-020-0736-7

Mostert-Kerckhoffs, M. A. L., Staal, W. G., Houben, R. H., & de Jonge, M. V. (2015). Stop and change: inhibition and flexibility skills are related to repetitive behavior in children and young adults with autism spectrum disorders. Journal of Autism and Developmental Disorders, 45(10), 3148–3158. doi:10.1007/s10803-015-2473-y

Mulder, M. J., Bos, D., Weusten, J. M. H., van Belle, J., van Dijk, S. C., Simen, P., … Durston, S. (2010). Basic impairments in regulating the speed-accuracy tradeoff predict symptoms of attention-deficit/hyperactivity disorder. Biological Psychiatry, 68(12), 1114–1119. doi:10.1016/j.biopsych.2010.07.031

Nader, A.-M., Jelenic, P., & Soulières, I. (2015). Discrepancy between WISC-III and WISC-IV Cognitive Profile in Autism Spectrum: What Does It Reveal about Autistic Cognition? PloS One, 10(12), e0144645. doi:10.1371/journal.pone.0144645

O’Hora, K. P., Lin, A., Kushan-Wells, L., & Bearden, C. E. (2022). Copy number variation at the 22q11.2 locus influences prevalence, severity, and psychiatric impact of sleep disturbance. Journal of Neurodevelopmental Disorders, 14(1), 41. doi:10.1186/s11689-022-09450-0

O’Hora, K. P., Zhang, Z., Vajdi, A., Kushan-Wells, L., Huang, Z. S., Pacheco-Hansen, L., … Bearden, C. E. (2022). Neurobehavioral Dimensions of Prader Willi Syndrome: Relationships Between Sleep and Psychosis-Risk Symptoms. Frontiers in Psychiatry / Frontiers Research Foundation, 13, 868536. doi:10.3389/fpsyt.2022.868536

Oliveras-Rentas, R. E., Kenworthy, L., Roberson, R. B., 3rd, Martin, A., & Wallace, G. L. (2012). WISC-IV profile in high-functioning autism spectrum disorders: impaired processing speed is associated with increased autism communication symptoms and decreased adaptive communication abilities. Journal of Autism and Developmental Disorders, 42(5), 655–664. doi:10.1007/s10803-011-1289-7

Olsen, L., Sparsø, T., Weinsheimer, S. M., Dos Santos, M. B. Q., Mazin, W., Rosengren, A., … Werge, T. (2018). Prevalence of rearrangements in the 22q11.2 region and population-based risk of neuropsychiatric and developmental disorders in a Danish population: a case-cohort study. The Lancet Psychiatry, 5(7), 573–580. doi:10.1016/S2215-0366(18)30168-8

Overall, J. E., & Gorham, D. R. (1962). The Brief Psychiatric Rating Scale. Psychological Reports, 10, 799–812. doi:10.2466/pr0.1962.10.3.799

Pinheiro, J., Bates, D., & R Core Team. (2022). nlme: Linear and Nonlinear Mixed Effects Models. Retrieved from https://CRAN.R-project.org/package=nlme

R Core Team. (2021). R: A Language and Environment for Statistical Computing. R Foundation for Statistical Computing.

Rees, E., Kirov, G., Sanders, A., Walters, J. T. R., Chambert, K. D., Shi, J., … Owen, M. J. (2014). Evidence that duplications of 22q11.2 protect against schizophrenia. Molecular Psychiatry, 19(1), 37–40. doi:10.1038/mp.2013.156

Roncone, R., Ventura, J., Impallomeni, M., Falloon, I. R., Morosini, P. L., Chiaravalle, E., & Casacchia, M. (1999). Reliability of an Italian standardized and expanded Brief Psychiatric Rating Scale (BPRS 4.0) in raters with high vs. low clinical experience. Acta Psychiatrica Scandinavica, 100(3), 229–236. doi:10.1111/j.1600-0447.1999.tb10850.x

Schneider, M., Debbané, M., Bassett, A. S., Chow, E. W. C., Fung, W. L. A., Van Den Bree, M. B. M., … Eliez, S. (2014). Psychiatric disorders from childhood to adulthood in 22q11.2 deletion syndrome: Results from the international consortium on brain and behavior in 22q11.2 deletion syndrome. The American Journal of Psychiatry, 171(6), 627–639. doi:10.1176/appi.ajp.2013.13070864

Service, S. K., Vargas Upegui, C., Castaño Ramírez, M., Port, A. M., Moore, T. M., Munoz Umanes, M., … Freimer, N. B. (2020). Distinct and shared contributions of diagnosis and symptom domains to cognitive performance in severe mental illness in the Paisa population: a case-control study. The Lancet. Psychiatry, 7(5), 411–419. doi:10.1016/S2215-0366(20)30098-5

Solomon, M., Gordon, A., Iosif, A.-M., Geddert, R., Krug, M. K., Mundy, P., & Hessl, D. (2021). Using the NIH Toolbox to Assess Cognition in Adolescents and Young Adults with Autism Spectrum Disorders. Autism Research: Official Journal of the International Society for Autism Research, 14(3), 500–511. doi:10.1002/aur.2399

Sønderby, I. E., Ching, C. R. K., Thomopoulos, S. I., van der Meer, D., Sun, D., Villalon-Reina, J. E., … ENIGMA 22q11.2 Deletion Syndrome Working Group. (2022). Effects of copy number variations on brain structure and risk for psychiatric illness: Large-scale studies from the ENIGMA working groups on CNVs. Human Brain Mapping, 43(1), 300–328. doi:10.1002/hbm.25354

Suzuki, G., Harper, K. M., Hiramoto, T., Funke, B., Lee, M., Kang, G., … Hiroi, N. (2009). Over-expression of a human chromosome 22q11.2 segment including TXNRD2, COMT and ARVCF developmentally affects incentive learning and working memory in mice. Human Molecular Genetics, 18(20), 3914–3925. doi:10.1093/hmg/ddp334

Venables, W. N., & Ripley, B. D. (2002). Modern Applied Statistics with S (Fourth). Fourth. Retrieved from https://www.stats.ox.ac.uk/pub/MASS4/

Verbesselt, J., Solot, C. B., Van Den Heuvel, E., Crowley, T. B., Giunta, V., Breckpot, J., … Swillen, A. (2023). Language Profiles of School-Aged Children with 22q11.2 Copy Number Variants. Genes, 14(3), 679. doi:10.3390/genes14030679

Verbesselt, J., Zink, I., Breckpot, J., & Swillen, A. (2022). Cross-sectional and longitudinal findings in patients with proximal 22q11.2 duplication: A retrospective chart study. American Journal of Medical Genetics. Part A, 188(1), 46–57. doi:10.1002/ajmg.a.62487

Vorstman, J. A. S., Breetvelt, E. J., Duijff, S. N., Eliez, S., Schneider, M., Jalbrzikowski, M., … International Consortium on Brain and Behavior in 22q11.2 Deletion Syndrome. (2015). Cognitive decline preceding the onset of psychosis in patients with 22q11.2 deletion syndrome. JAMA Psychiatry, 72(4), 377–385. doi:10.1001/jamapsychiatry.2014.2671

Wechsler, D. (1999). Manual for the Wechsler Abbreviated Intelligence Scale (WASI). San Antonio, TX: The Psychological Corporation.

Wechsler, David. (n.d.). Wechsler Abbreviated Scale of Intelligence. doi:10.1037/t15170-000

Weinberger, R., Yi, J., Calkins, M., Guri, Y., McDonald-McGinn, D. M., Emanuel, B. S., … Gothelf, D. (2016). Neurocognitive profile in psychotic versus nonpsychotic individuals with 22q11.2 deletion syndrome. European Neuropsychopharmacology: The Journal of the European College of Neuropsychopharmacology, 26(10), 1610–1618. doi:10.1016/j.euroneuro.2016.08.003

Wickham, H. (2016). ggplot2: Elegant Graphics for Data Analysis. Springer-Verlag New York.

Yi, J. J., Weinberger, R., Moore, T. M., Calkins, M. E., Guri, Y., McDonald-McGinn, D. M., … Gur, R. C. (2016). Performance on a computerized neurocognitive battery in 22q11.2 deletion syndrome: A comparison between US and Israeli cohorts. Brain and Cognition, 106, 33–41. doi:10.1016/j.bandc.2016.02.002

