## Supplementary Materials for "Distinct Neurocognitive Profiles and Clinical Phenotypes Associated with Copy Number Variation at the 22q11.2 Locus"

**Table S1.** Characteristics of 22q11.2 CNV Carriers with Autism Diagnostic data Available (N=68)

|  | 22qDelASD- | 22qDelASD+ | 22qDupASD- | 22qDupASD+ |
| --- | --- | --- | --- | --- |
| *n* | 22 | 18 | 17 | 11 |
| Age, Years (SD) | 19.9 (10.4) | 14.8 (4.4) | 17.8 (14.7) | 11.6 (3.3) |
| Age Range, Years | 7-41 | 9-24 | 6-49 | 7-17 |
| Males, *n* (%) | 8 (36.4%) | 8 (44.4%) | 9 (52.9%) | 7 (63.6%) |
| Psychotic Spectrum Symptoms^a^, *n* (%) | 5 (27.8%) | 8 (47.1%) | 2 (16.2%) | 4 (37.5%) |
| ADHD, *n* (%) | 8 (36.4%) | 13 (72.2%) | 6 (35.3%) | 7 (63.6%) |
| Anxiety Disorder, *n* (%) | 10 (45.5%) | 12 (66.7%) | 7 (41.2%) | 8 (72.7%) |

**^a^**Any positive symptom rated >2 on the SIPS

**Table S2.** Results of LMMS comparing Neurocognitive Profiles

|  | Group*variable Interaction | | Group*Age Interaction | | Group*Variable*Age Interaction | |
| --- | --- | --- | --- | --- | --- | --- |
|  | $\chi^{2}$ | *p* | $\chi^{2}$ | *p* | $\chi^{2}$ | *p* |
| Accuracy | 51.32 | **<0.001** | 6.982 | **0.0305** | 13.314 | 0.924 |
| Speed | 88.82 | **<0.001** | 2.361 | 0.307 | 97.506 | **0.010** |
| Accuracy + Anti-psychotic use | 38.80 | **0.015** | 3.906 | 0.142 | 10.057 | 0.986 |
| Speed + Anti-psychotic use | 59.42 | **<0.001** | 2.23 | 0.327 | 19.054 | 0.834 |

**Table S3.** Significant Group-by-Age Relationships

| **Variable** | **Comparison** | ***b*** | ***p*** |
| --- | --- | --- | --- |
| Emotion Differentiation | TD>22qDel | -0.726 | 0.009 |
| Emotion Differentiation | TD>22qDup | -0.597 | 0.022 |
| Verbal Reasoning | TD>22qDel | -0.68 | 0.016 |
| Mental Flexibility | TD>22qDel | -0.692 | 0.012 |
| Mental Flexibility | TD>22qDup | -0.514 | 0.047 |
| Age Differentiation | TD>22qDel | -0.761 | 0.006 |
| Age Differentiation | TD>22qDup | -0.63 | 0.014 |
| Sensorimotor Speed | 22qDel>22qDup | 0.464 | 0.046 |

**Table S4.** Summary of LMMs Assessing Between Group Differences in Penn-CNB Domains

|  | **TD>22qDel** | | | | **TD>22qDup** | | | | **22qDel > 22qDup** | | | |
| --- | --- | --- | --- | --- | --- | --- | --- | --- | --- | --- | --- | --- |
|  | ***b*** | ***95% CI*** | ***p*** | ***q*** | ***b*** | ***95% CI*** | ***p*** | ***q*** | ***b*** | ***95% CI*** | ***p*** | ***q*** |
| **Accuracy** | | | | | | | | | | | | |
| Complex Cognition | **−1.45** | **-1.81,**  **-1.09** | **<0.001** | **<0.001** | **−1.11** | **-1.50,**  **-0.72** | **<0.001** | **<0.001** | 0.35 | -0.02, 0.706 | 0.068 | 0.136 |
| Episodic Memory | **−1.45** | **-1.79,**  **-1.10** | **<0.001** | **<0.001** | **−0.91** | **-1.29,**  **-0.52** | **<0.001** | **<0.001** | **0.54** | **0.20, 0.88** | **0.003** | **0.012** |
| Executive Control | **−1.02** | **-1.36,**  **-0.69** | **<0.001** | **<0.001** | **−0.89** | **-1.27,**  **-0.51** | **<0.001** | **<0.001** | 0.13 | -0.23, 0.49 | 0.492 | 0.656 |
| Social Cognition | **−1.19** | **-1.54,**  **-0.83** | **<0.001** | **<0.001** | **−1.19** | **-1.58,**  **-0.79** | **<0.001** | **<0.001** | −0.00 | -0.35, 0.34 | 0.993 | 0.993 |
| **Speed** | | | | | | | | | | | | |
| Complex Cognition | **−0.63** | **-1.06,**  **-0.19** | **0.007** | **0.017** | **−0.77** | **-1.24,**  **-0.29** | **0.002** | **0.004** | −0.14 | -0.58, 0.30 | 0.543 | 0.543 |
| Episodic Memory | −0.18 | -0.60, 0.24 | 0.403 | 0.504 | **−0.62** | **-1.08,**  **-0.16** | **0.010** | **0.010^*^** | −0.44 | -0.86,  -0.04 | 0.038^*^ | 0.063 |
| Executive Control | −0.10 | -0.49, 0.30 | 0.641 | 0.641 | −0.71 | -1.16,  -0.26 | 0.003 | 0.004 | **−0.62** | **-1.05,**  **-0.18** | **0.007** | **0.017** |
| Sensorimotor | **−0.41** | **-0.74,**  **-0.10** | **0.015** | **0.025** | **−1.20** | **-1.58,**  **-0.82** | **<0.001** | **<0.001** | **−0.79** | **-1.14, -0.43** | **<0.001** | **<0.001** |
| Social Cognition | **−0.63** | **-1.03,**  **-0.23** | **0.003** | **0.015** | **−0.86** | **-1.31,**  **-0.41** | **<0.001** | **0.001** | −0.22 | -0.62, 0.16 | 0.260 | 0.326 |
| CI: Confidence Interval | | | | | | | | | | | | |

**Table S5.** Summary of LMMs Assessing Between Group Differences in Individual Penn-CNB Tests

|  | **TD>22qDel** | | | | **TD>22qDup** | | | | **22qDel > 22qDup** | | | |
| --- | --- | --- | --- | --- | --- | --- | --- | --- | --- | --- | --- | --- |
|  | ***b*** | ***95% CI*** | ***p*** | ***q*** | ***b*** | ***95% CI*** | ***p*** | ***q*** | ***b*** | ***95% CI*** | ***p*** | ***q*** |
| **Accuracy** | | | | | | | | | | | | |
| Age Differentiation | **−1.17** | **-1.48, -0.86** | **<0.001** | **<0.001** | **−1.18** | **-1.53, -0.83** | **<0.001** | **<0.001** | −0.01 | -0.33, 0.32 | 0.964 | 0.966 |
| Face Memory | **−1.57** | **-1.86, -1.29** | **<0.001** | **<0.001** | **−1.01** | **-1.33, -0.68** | **<0.001** | **<0.001** | **0.57** | **0.27, 0.87** | **<0.001** | **0.004** |
| Emotion Identification | **−1.05** | **-1.38, -0.72** | **<0.001** | **<0.001** | **−1.05** | **-1.42, -0.67** | **<0.001** | **<0.001** | 0.01 | -0.34, 0.36 | 0.966 | 0.966 |
| Word Memory | **−1.10** | **-1.47, -0.73** | **<0.001** | **<0.001** | **−0.76** | **-1.17, -0.35** | **0.001** | **0.001** | 0.34 | -0.02, 0.7 | 0.072 | 0.177 |
| Language Reasoning | **−1.34** | **-1.70, -0.99** | **<0.001** | **<0.001** | **−1.02** | **-1.40, -0.63** | **<0.001** | **<0.001** | 0.33 | -0.02, 0.68 | 0.074 | 0.177 |
| Spatial Ability | **−1.15** | **-1.47, -0.82** | **<0.001** | **<0.001** | **−1.01** | **-1.37, -0.64** | **<0.001** | **<0.001** | 0.14 | -0.21, 0.49 | 0.445 | 0.715 |
| Emotion Differentiation | **−1.18** | **-1.56, -0.81** | **<0.001** | **<0.001** | **−1.06** | **-1.48, -0.64** | **<0.001** | **<0.001** | 0.12 | -0.24, 0.48 | 0.517 | 0.715 |
| Mental Flexibility | **−1.03** | **-1.37, -0.68** | **<0.001** | **<0.001** | **−0.65** | **-1.04, -0.27** | **0.001^*^** | **0.001^*^** | 0.373 | 0.01, 0.73 | **0.048** | 0.177 |
| Nonverbal Reasoning | **−1.57** | **-1.88, -1.26** | **<0.001** | **<0.001** | **−0.98** | **-1.33, -0.63** | **<0.001** | **<0.001** | **0.590** | **0.26, 0.92** | **0.001** | **0.005** |
| Working Memory | **−0.78** | **-1.11, -0.44** | **<0.001** | **<0.001** | **−0.76** | **-1.14, -0.38** | **<0.001** | **<0.001** | 0.02 | -0.34, 0.37 | 0.923 | 0.966 |
| Attention | **−0.53** | **-0.85, -0.20** | **0.002** | **0.002** | **−0.63** | **-0.10, -0.27** | **0.001** | **0.001** | −0.11 | -0.43, 0.22 | 0.537 | 0.715 |
| Spatial Memory | **−0.97** | **-1.30, -0.64** | **<0.001** | **<0.001** | **−0.68** | **-1.06, -0.31** | **0.001** | **0.001** | 0.29 | -0.06, 0.63 | 0.108 | 0.215 |
| **Speed** |  |  |  |  |  |  |  |  |  |  |  |  |
| Age Differentiation | **−0.57** | **-0.93, -0.22** | **0.002^*^** | **0.009^*^** | **−0.67** | **-1.08, -0.27** | **0.002** | **0.004** | −0.10 | -0.47, 0.28 | 0.613 | 0.660 |
| Face Memory | −0.22 | -0.59, 0.16 | 0.267 | 0.339 | −0.46 | -0.88, -0.03 | 0.041^*^ | 0.058 | −0.24 | -0.63, 0.16 | 0.243 | 0.416 |
| Emotion Identification | **−0.79** | **-1.14, -0.44** | **<0.001** | **<0.001** | **−0.73** | **-1.14, -0.33** | **0.001** | **0.002** | 0.057 | -0.32, 0.43 | 0.769 | 0.769 |
| Word Memory | −0.28 | -0.68, 0.13 | 0.189 | 0.265 | **−0.50** | **-0.94, -0.06** | **0.031** | **0.048** | −0.23 | -0.62, 0.17 | 0.268 | 0.416 |
| Language Reasoning | **−0.55** | **-0.94, -0.15** | **0.009** | **0.024** | −0.42 | -0.85, 0.01 | 0.061 | 0.077 | 0.12 | -0.26, 0.50 | 0.534 | 0.628 |
| Spatial Ability | **−1.12** | **-1.46, -0.79** | **<0.001** | **<0.001** | **−0.97** | **-1.34, -0.60** | **<0.001** | **<0.001** | 0.15 | -0.21, 0.520 | 0.422 | 0.591 |
| Emotion Differentiation | −0.33 | -0.72, 0.051 | 0.097 | 0.150 | **−0.65** | **-1.08, -0.22** | **0.004** | **0.008** | −0.31 | -0.68, 0.05 | 0.101 | 0.284 |
| Sensorimotor Processing Speed | −0.35 | -0.70, -0.01 | 0.052 | 0.092 | **−0.89** | **-1.28, -0.49** | **<0.001** | **<0.001** | **−0.53** | **-0.90, -0.16** | **0.007** | **0.031** |
| Mental Flexibility | −0.44 | -0.82, -0.07 | 0.026^*^ | 0.051 | **−0.57** | **-1.00, -0.14** | **0.012** | **0.021** | −0.13 | -0.53, 0.27 | 0.538 | 0.628 |
| Nonverbal Reasoning | **0.77** | **0.39, 1.15** | **<0.001** | **0.001** | 0.21 | -0.22, 0.65 | 0.350 | 0.350 | **−0.56** | **-0.97, -0.15** | **0.010** | **0.035** |
| Motor Speed | **−0.39** | **-0.71, -0.07** | **0.020** | **0.047** | **−1.26** | **-1.64, -0.89** | **<0.001** | **<0.001** | **−0.87** | **-1.22, -0.53** | **<0.001** | **<0.001** |
| Working Memory | 0.03 | -0.36, 0.41 | 0.892 | 0.904 | −0.31 | -0.75, 0.13 | 0.174 | 0.203 | −0.34 | -0.76, 0.08 | 0.122 | 0.284 |
| Attention | −0.03 | -0.39, 0.33 | 0.874 | 0.904 | **−0.71** | **-1.11, -0.32** | **0.001** | **0.002** | **−0.68** | **-1.06, -0.30** | **0.001** | **0.005** |
| Spatial Memory | 0.02 | -0.35, 0.39 | 0.904 | 0.904 | −0.24 | -0.66, 0.18 | 0.264 | 0.285 | −0.27 | -0.66, 0.12 | 0.188 | 0.376 |
| CI: Confidence Interval | | | | | | | | | | | | |

**Table S6.** Summary of LMMs Assessing Associations between Penn-CNB Domains and Clinical Outcomes

|  | **Within 22qDel** | | | | **Interaction** | | | | **Within 22qDup** | | | |
| --- | --- | --- | --- | --- | --- | --- | --- | --- | --- | --- | --- | --- |
|  | ***b*** | ***95% CI*** | ***p*** | ***q*** | ***b*** | ***95% CI*** | ***p*** | ***q*** | ***b*** | ***95% CI*** | ***p*** | ***q*** |
| **Accuracy- Global Psychopathology** | | | | | | | | | | | | |
| Complex Cognition | −0.09 | -0.24, 0.06 | 0.25 | 0.42 | −0.01 | -0.33, 0.30 | 0.95 | 0.95 | −0.10 | -0.39, 0.18 | 0.50 | 0.66 |
| Episodic Memory | 0.01 | -0.18, 0.20 | 0.91 | 0.91 | −0.23 | -0.57, 0.12 | 0.20 | 0.52 | −0.22 | -0.51, 0.07 | 0.15 | 0.30 |
| Executive Control | −0.10 | -0.30, 0.09 | 0.31 | 0.42 | −0.18 | -0.54, 0.17 | 0.33 | 0.52 | −0.29 | -0.59, 0.01 | 0.07 | 0.29 |
| Social Cognition | −0.17 | -0.34, 0.00 | 0.06 | 0.25 | 0.14 | -0.19, 0.46 | 0.39 | 0.52 | −0.02 | -0.31, 0.25 | 0.88 | 0.88 |
| **Accuracy – Social Problems** | | | | | | | | | | | | |
| Complex Cognition | 0.05 | -0.18, 0.28 | 0.66 | 0.66 | −0.18 | -0.49, 0.14 | 0.28 | 0.57 | −0.13 | -0.33, 0.08 | 0.25 | 0.25 |
| Episodic Memory | 0.23 | -0.12, 0.56 | 0.20 | 0.40 | −0.48 | -0.96, 0.04 | 0.07 | 0.29 | −0.25 | -0.58, 0.11 | 0.18 | 0.24 |
| Executive Control | −0.29 | -0.63, 0.07 | 0.12 | 0.40 | −0.16 | -0.64, 0.33 | 0.54 | 0.67 | −0.45 | -0.78, -0.07 | **0.014** | 0.05 |
| Social Cognition | −0.11 | -0.37, 0.13 | 0.42 | 0.56 | −0.08 | -0.43, 0.28 | 0.67 | 0.67 | −0.19 | -0.44, 0.05 | 0.16 | 0.24 |
| **Accuracy – Global Psychosocial Functioning** | | | | | | | | | | | | |
| Complex Cognition | 0.07 | -0.09, 0.24 | 0.38 | 0.38 | −0.37 | -0.66, -0.07 | **0.018** | 0.07 | −0.30 | -0.55, -0.02 | **0.025** | 0.10 |
| Episodic Memory | 0.19 | -0.00, 0.39 | 0.06 | 0.23 | −0.26 | -0.61, 0.10 | 0.15 | 0.30 | −0.07 | -0.36, 0.24 | 0.65 | 0.83 |
| Executive Control | 0.18 | -0.04, 0.41 | 0.12 | 0.23 | −0.15 | -0.53, 0.25 | 0.47 | 0.47 | 0.04 | -0.28, 0.38 | 0.83 | 0.83 |
| Social Cognition | 0.13 | -0.06, 0.32 | 0.19 | 0.25 | −0.16 | -0.47, 0.15 | 0.31 | 0.41 | −0.04 | -0.29, 0.22 | 0.77 | 0.83 |
| **Accuracy- Positive Symptoms** | | | | | | | | | | | | |
| Complex Cognition | −0.06 | -0.21, 0.09 | 0.42 | 0.48 | −0.05 | -0.43, 0.32 | 0.79 | 0.97 | −0.11 | -0.46, 0.23 | 0.52 | 0.69 |
| Episodic Memory | −0.06 | -0.23, 0.11 | 0.48 | 0.48 | 0.25 | -0.22, 0.71 | 0.31 | 0.97 | 0.19 | -0.25, 0.63 | 0.41 | 0.69 |
| Executive Control | −0.08 | -0.26, 0.10 | 0.39 | 0.48 | −0.01 | -0.53, 0.49 | 0.97 | 0.97 | −0.09 | -0.57, 0.38 | 0.71 | 0.71 |
| Social Cognition | −0.12 | -0.28, 0.046 | 0.16 | 0.48 | −0.05 | -0.48, 0.39 | 0.83 | 0.97 | −0.16 | -0.56, 0.25 | 0.43 | 0.69 |
| **Speed- Global Psychopathology** | | | | | | | | | | | | |
| Complex Cognition | −0.14 | -0.41, 0.10 | 0.28 | 0.69 | 0.20 | -0.39, 0.78 | 0.52 | 0.65 | 0.06 | -0.48, 0.58 | 0.84 | 0.84 |
| Episodic Memory | 0.00 | -0.18, 0.19 | 1.00 | 1.00 | −0.16 | -0.50, 0.19 | 0.37 | 0.62 | −0.16 | -0.45, 0.14 | 0.29 | 0.48 |
| Executive Control | 0.01 | -0.15, 0.17 | 0.86 | 1.00 | −0.27 | -0.58, 0.05 | 0.10 | 0.24 | −0.26 | -0.52, 0.01 | 0.07 | 0.17 |
| Sensorimotor | −0.10 | -0.27, 0.07 | 0.24 | 0.69 | 0.03 | -0.29, 0.34 | 0.86 | 0.86 | −0.07 | -0.34, 0.19 | 0.60 | 0.75 |
| Social Cognition | 0.04 | -0.15, 0.24 | 0.68 | 1.00 | **−0.55** | **-0.91, -0.18** | **0.004** | **0.021** | **−0.51** | **-0.82, -0.19** | **0.002** | **0.009** |
| **Speed- Social Problems** | | | | | | | | | | | | |
| Complex Cognition | −0.11 | -0.49, 0.28 | 0.60 | 0.92 | −0.44 | -1.05, 0.16 | 0.19 | 0.24 | **−0.55** | **-0.99, -0.12** | **0.025** | **0.031** |
| Episodic Memory | −0.16 | -0.52, 0.22 | 0.43 | 0.92 | −0.52 | -1.08, 0.02 | 0.08 | 0.13 | **−0.68** | **-1.06, -0.30** | **0.002** | **0.004** |
| Executive Control | −0.06 | -0.40, 0.27 | 0.74 | 0.92 | −0.04 | -0.72, 0.46 | 0.89 | 0.89 | −0.10 | -0.67, 0.29 | 0.57 | 0.57 |
| Sensorimotor | 0.23 | -0.10, 0.53 | 0.15 | 0.77 | **−0.70** | **-1.13, -0.26** | **0.004** | **0.010** | **−0.48** | **-0.78, -0.18** | **0.005** | **0.008** |
| Social Cognition | 0.00 | -0.31, 0.30 | 1.00 | 1.00 | **−0.73** | **-1.18, -0.29** | **0.004** | **0.010** | **−0.73** | **-1.06, -0.42** | **<.001** | **<.001** |
| **Speed- Global Psychosocial Functioning** | | | | | | | | | | | | |
| Complex Cognition | 0.26 | 0.01, 0.53 | 0.06 | 0.29 | −0.08 | -0.57, 0.41 | 0.76 | 0.76 | 0.18 | -0.23, 0.61 | 0.41 | 0.51 |
| Episodic Memory | −0.15 | -0.36, 0.06 | 0.17 | 0.42 | 0.41 | 0.03, 0.79 | 0.037^*^ | 0.09 | 0.26 | -0.07, 0.58 | 0.12 | 0.29 |
| Executive Control | 0.02 | -0.17, 0.21 | 0.83 | 0.83 | 0.16 | -0.16, 0.48 | 0.33 | 0.41 | 0.18 | -0.08, 0.45 | 0.18 | 0.29 |
| Sensorimotor | 0.10 | -0.08, 0.27 | 0.30 | 0.50 | −0.19 | -0.50, 0.15 | 0.26 | 0.41 | −0.09 | -0.36, 0.19 | 0.52 | 0.52 |
| Social Cognition | −0.03 | -0.25, 0.18 | 0.79 | 0.83 | 0.45 | 0.08, 0.82 | 0.020^*^ | 0.09 | **0.42** | **0.11, 0.72** | **0.008^*^** | **0.042^*^** |
| **Speed- Positive Psychosis-risk Symptoms** | | | | | | | | | | | | |
| Complex Cognition | 0.00 | -0.22, 0.22 | 0.99 | 0.99 | 0.22 | -0.49, 1.02 | 0.55 | 0.64 | 0.22 | -0.44, 0.96 | 0.53 | 0.67 |
| Episodic Memory | 0.02 | -0.15, 0.19 | 0.83 | 0.99 | −0.17 | -0.69, 0.36 | 0.51 | 0.64 | −0.158 | -0.64, 0.34 | 0.54 | 0.67 |
| Executive Control | 0.01 | -0.14, 0.16 | 0.89 | 0.99 | −0.32 | -0.72, 0.08 | 0.12 | 0.30 | −0.309 | -0.68, 0.06 | 0.10 | 0.28 |
| Sensorimotor | −0.06 | -0.2, 0.09 | 0.47 | 0.99 | 0.11 | -0.33, 0.54 | 0.64 | 0.64 | 0.05 | -0.36, 0.46 | 0.81 | 0.81 |
| Social Cognition | 0.03 | -0.16, 0.214 | 0.78 | 0.99 | −0.44 | -0.96, 0.09 | 0.11 | 0.30 | −0.409 | -0.9, 0.09 | 0.11 | 0.28 |
| CI: Confidence Interval | | | | | | | | | | | | |

**Table S7.** Main Effects of LMMs Comparing Overall Neurocognitive Profiles between 22q11.2 CNV Carriers with and without ASD

|  | Group*variable Interaction | | Group*Age Interaction | | Group*Variable*Age Interaction | |
| --- | --- | --- | --- | --- | --- | --- |
|  | $\chi^{2}$ | *p* | $\chi^{2}$ | *p* | $\chi^{2}$ | *p* |
| Accuracy | 43.5 | 0.104 | 7.869 | **0.0488** | 52.60 | **0.017** |
| Speed | 94.37 | **<0.001** | 2.57 | 0.463 | 47.15 | 0.173 |

**Table S8.** Summary of LMMs Assessing Differences in Penn-CNB Domains between 22q11.2 CNV Carriers with and without ASD

|  | **Executive Control** | | | **Episodic Memory** | | | **Complex Cognition** | | | **Social Cognition** | | | | **Sensorimotor** | | | |
| --- | --- | --- | --- | --- | --- | --- | --- | --- | --- | --- | --- | --- | --- | --- | --- | --- | --- |
|  | ***b*** | ***95% CI*** | ***q*** | ***b*** | ***95% CI*** | ***q*** | ***b*** | ***95% CI*** | ***q*** | | ***b*** | ***95% CI*** | ***q*** | | ***b*** | ***95% CI*** | ***q*** |
| 22qDelASD+ > 22qDelASD- | -0.08 | -0.66,  0.51 | 0.96 | -0.59 | -1.12, -0.07 | 0.165 | -0.02 | -0.65, 0.62 | 0.96 | | -0.25 | -0.85, 0.32 | 0.96 | | -0.11 | -0.56, 0.34 | 0.96 |
| 22qDelASD+ > 22qDupASD- | -0.87 | -1.50, -0.22 | **0.021** | -0.91 | -1.47, -0.36 | **<0.001** | -0.22 | -0.87, 0.43 | 0.529 | | -0.63 | -1.24, -0.02 | 0.067 | | -0.94 | -1.44, -0.45 | **0.003** |
| 22qDelASD+ > 22qDupASD+ | -1.00 | -1.73, -0.27 | **0.029** | -0.34 | -0.98, 0.30 | 0.524 | 0.22 | -0.53, 0.97 | 0.577 | | -0.27 | -0.96, 0.41 | 0.569 | | -0.90 | -1.46, -0.34 | **0.015** |
| 22qDelASD- > 22qDupASD- | -0.79 | -1.40, -0.169 | **0.048** | -0.32 | -0.86, 0.23 | 0.34 | -0.20 | -0.84, 0.44 | 0.56 | | -0.38 | -0.98, 0.23 | 0.34 | | -0.83 | -1.31, -0.34 | **0.008** |
| 22qDelASD- > 22qDupASD+ | -0.93 | -1.68, -0.17 | 0.059 | 0.26 | -0.40, 0.91 | 0.707 | 0.24 | -0.55, 1.03 | 0.707 | | -0.02 | -0.72, 0.69 | 0.96 | | -0.78 | -1.35, -0.21 | **0.05** |
| 22qDupASD- > 22qDupASD+ | -0.14 | -0.90, 0.62 | 0.878 | 0.58 | -0.06, 1.22 | 0.451 | 0.44 | -0.33, 1.22 | 0.514 | | 0.36 | -0.32, 1.04 | 0.514 | | 0.05 | -0.52, 0.60 | 0.878 |

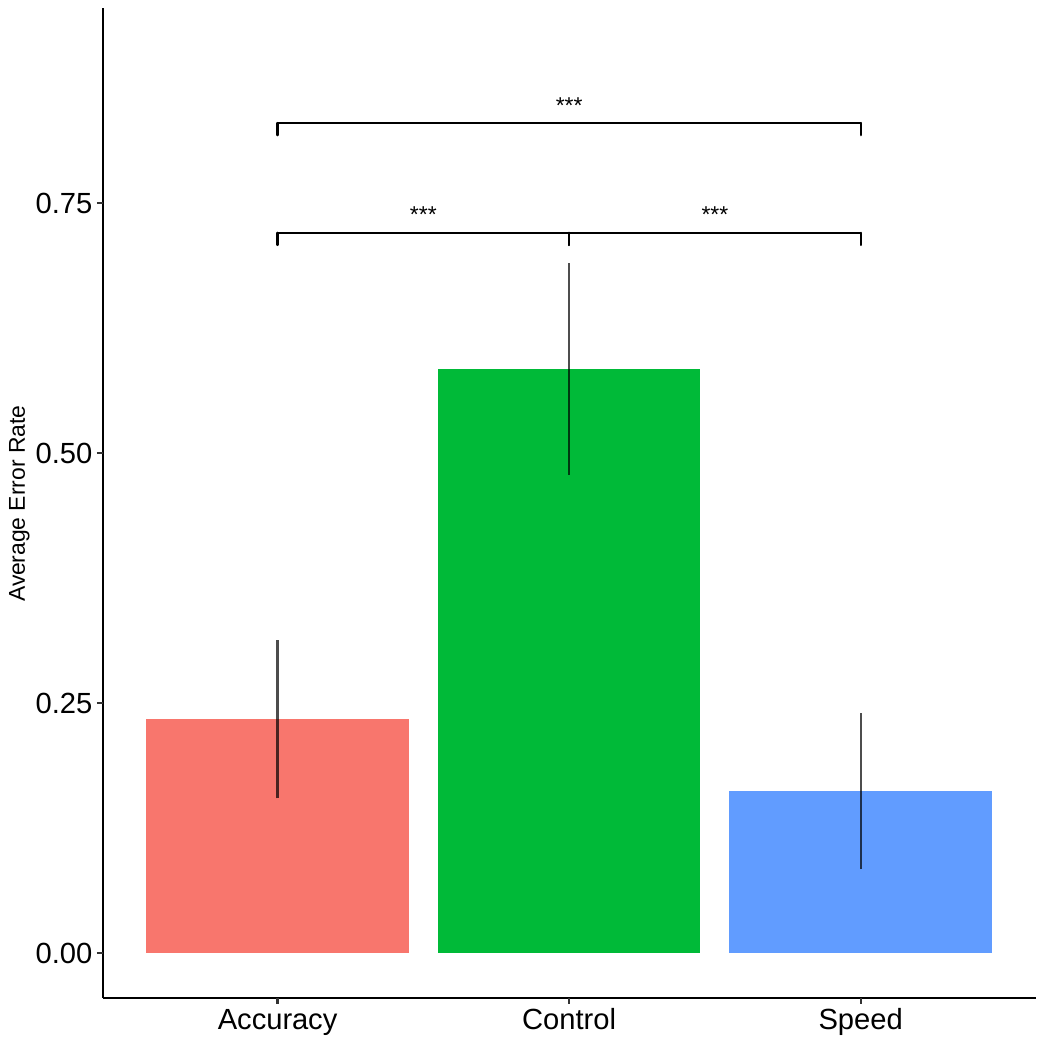

**Figure S1.** Discrimination accuracy (mean ± standard deviation) across 100 trials for the two LDA models differentiating 22qDel Carriers from 22qDup carriers (accuracy only: red; speed only: blue) and the control model (green). Both models performed significantly better than the control model, but the Speed model was significantly more accurate than the accuracy model. ^*^*q*<0.05, ^**^*q*<0.01, ^***^*q*<0.001
